# Resolving Parameter Uncertainty in Outbreak Models Through Population-Level Serological Surveillance

**DOI:** 10.1101/2025.10.09.25337678

**Authors:** Binod Pant, Matthew E. Levine, Anjalika Nande, Raúl Garrido García, George Dewey, Nicholas B. Link, Mauricio Santillana

**Affiliations:** Machine Intelligence Group for the Betterment of Health and the Environment, Northeastern University, Boston, MA, 02115, USA; Network Science Institute, Northeastern University, Boston, MA, 02115, USA; Eric and Wendy Schmidt Center, Broad Institute of MIT and Harvard, Cambridge, MA, 02142, USA; Basis Research Institute, Cambridge, MA, 02142, USA; Institute for Computational Medicine, Johns Hopkins University, Baltimore, MD, 21218, USA; Department of Physics, Northeastern University, Boston, MA, 02115, USA; Department of Biostatistics, Harvard T.H. Chan School of Public Health, Boston, MA, 02115, USA; Department of Epidemiology, Harvard T.H. Chan School of Public Health, Boston, MA, 02115, USA

**Keywords:** Uncertainty quantification, Parameter uncertainty, Seroprevalence data, Bayesian inference

## Abstract

Epidemic models face a critical challenge: surveillance systems capture only a fraction of infections (often *<* 10%). We reveal two fundamental problems. First, when models ignore underdetection entirely—treating detected cases as complete—parameter errors exceed 1000% despite visually reasonable fits. Second, when models explicitly account for underdetection by including case detection ratios as unknown parameters, structural identifiability analysis proves transmission rates and detection ratios become mathematically confounded—rendering infinite epidemiologically distinct scenarios equally plausible from case data alone. Integrating even a single population-level seroprevalence measurement resolves both problems by independently constraining cumulative exposure. Through Bayesian inference on synthetic SIR data, we demonstrate that this approach reduces parameter uncertainty by orders of magnitude, enabling accurate inference of transmission dynamics, peak timing, and outbreak size under realistic noise. Our framework establishes serological surveillance integration as both a mathematical necessity and a strategic investment for pandemic preparedness.

## 1 Introduction

Across infectious diseases—from H1N1 influenza to dengue fever and COVID-19—surveillance systems consistently capture only a fraction of actual infections, with documented cases representing anywhere from 10% to less than 0.1% of true infection burden [1–8]. This underascertainment of infections produces a systematic bias which is further distorted by time-varying factors including testing fluctuations, reporting delays, and policy shifts. When mathematical models of infectious disease transmission are calibrated to directly match underascertained epidemiological case counts without acknowledging this underdetection problem [9–12], they generate severely biased—frequently by several orders of magnitude—parameter estimates that can differ dramatically from those likely occurring in the population during epidemic outbreaks. A model might suggest an outbreak is nearly contained when community transmission is actually accelerating, or recommend insufficient vaccine coverage when herd immunity thresholds are much higher than estimated.

The COVID-19 pandemic placed mathematical models at the forefront of critical public health decisionmaking affecting millions of lives and trillions in economic activity [13, 14]. A major challenge faced by modeling efforts was the sparsity and unreliability of early pandemic data—true infections worldwide were estimated to be 3 to 1000 times higher than those reported [15, 16]. The combination of poor data quality with many models’ limited ability to account for this massive underdetection led to seemingly accurate (model to data) fits but catastrophically biased estimates of transmission dynamics and intervention effectiveness that varied widely across models [17–19]. The epidemiological modeling community eventually recognized this limitation [20], which ultimately motivated modeling efforts such as the US COVID-19 Forecast Hub [21] to rethink the use of incidence case data as modeling targets.

The stakes of parameter misestimation are not merely theoretical. During the COVID-19 pandemic, forecasts from a variety of modeling approaches — including machine learning models and compartmental models — were used in state and national planning. However, retrospective evaluation of four widely used models revealed highly inaccurate point predictions and poor calibration of uncertainty (e.g. only 10% of predictions fell within 10% of the “ground truth,” and most 95% prediction intervals failed to achieve nominal coverage) [22]. This case highlights how misestimation of key epidemiological parameters - such as ascertainment ratio, effective reproduction numbers, infection rates, or epidemic size - can mislead scientific conclusions and risk leading to misguided policy decisions.

The mathematical roots of this crisis involve two distinct but related problems. First, when models ignore case underascertainment entirely—treating detected cases as if they represent all infections—the resulting parameter estimates become systematically biased, with errors that worsen as detection ratio decline. Even when such models achieve visually convincing fits to observed case data, their estimates of transmission rates, cumulative infections, recovery rates, and reproduction numbers can be catastrophically uninformative [20].

Second, when models explicitly account for incomplete detection by including case ascertainment ratios as unknown parameters, they face well-documented structural identifiability problems [17, 23]. The transmission rate and detection ratio become mathematically confounded in the observed data, meaning that multiple epidemiologically distinct scenarios can fit the data equally well. Standard inference procedures cannot distinguish between high transmission with low detection versus low transmission with high detection, creating what we term an “identifiability crisis” where confident-looking parameter estimates rest on mathematically unstable foundations.

Current approaches to address case underascertainment fall short of resolving these problems. Some studies attempt to correct for underdetection by incorporating hospitalization or mortality data [24, 25], but these approaches require accurate estimates of case-hospitalization or case-fatality ratios—themselves subject to the same ascertainment biases [26, 27]—in order to properly work [28]. Others explicitly model case detection processes by including case ascertainment ratios as unknown parameters [8, 29], but without additional constraining data sources, this approach may even worsen identifiability problems by introducing new unidentifiable parameters that cannot resolve the underlying bias. The result is a modeling field where models achieve good fits to observed case data, and thus produce seemingly precise epidemiological estimates, while harboring fundamental uncertainties that propagate through all derived quantities of interest, from basic reproduction numbers to epidemic peak timing and cumulative infections.

Here, we demonstrate that simultaneously fitting models to observed case data and population-level seroprevalence data, in the context of historical reconstructions of an epidemic outbreak, provides a mathematically rigorous solution to this modeling crisis. Unlike case surveillance, which captures ongoing infections imperfectly, seroprevalence surveys can directly measure cumulative population exposure when properly designed and sampled [30–32]. For models that ignore underascertainment entirely, seroprevalence data reveal the true scale of population exposure, allowing us to reject these fundamentally biased models. For models that explicitly include detection ratio as unknown parameters, seroprevalence measurements break the mathematical confounding between transmission rates and ascertainment ratio by providing an independent constraint on cumulative infections. By integrating even a single well-timed seroprevalence measurement with detected case data, the identifiability crisis dissolves: parameter uncertainties drop by orders of magnitude, enabling reliable inference of critical epidemic quantities including transmission rates, peak timing, and final outbreak size.

Through comprehensive computational (synthetic) experiments using SIR models [33], we demonstrate that this approach transforms unreliable inference into robust parameter estimation. Our analysis reveals that the benefits persist under realistic values of noise—with up to 40% measurement noise in underreported case counts and 10% in seroprevalence measurements—making the approach practically viable. Most importantly, we quantify the economic case for implementation, showing that population-level serosurvey costs are orders of magnitude smaller than the healthcare expenditures prevented through improved outbreak modeling.

Overall, our findings expose a systematic vulnerability in pandemic preparedness while providing an actionable remedy. As the world prepares for future pandemic threats, integrating serological surveillance into routine epidemiological modeling represents both a mathematical necessity and a strategic investment in more reliable outbreak response. More broadly, our uncertainty quantification framework provides a systematic approach for identifying when surveillance data are adequate for their intended modeling purpose—a critical capability for evidence-based public health decision making.

## 2 Mathematical Framework

### 2.1 The Parameter Identifiability Problem in Epidemiological Models

We demonstrate our computational approach using the canonical Susceptible-Infectious-Recovered (SIR) model [33], which serves as a prototype for a broad class of compartmental models facing similar identifiability challenges. The model categorizes the population into three compartments and evolves according to:

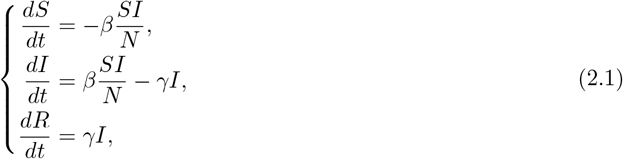

where *β* is the transmission rate, *γ* is the recovery rate, and *N* = *S* + *I* + *R* is the total population. While simple, this framework already showcases identifiability challenges that are present in more complex epidemiological models when the reality of undetected cases is considered.

The computational challenge emerges when surveillance systems observe only a fraction of true cases. If *I*_new_(*t*) = *βS*(*t*)*I*(*t*)*/N* represents true incidence, then detected incidence becomes:

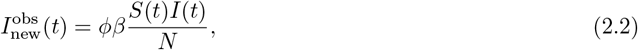

where *ϕ* ∈ (0, 1] is the case ascertainment ratio. This creates a fundamental inverse problem: multiple parameter combinations (*β, ϕ*) can produce identical observed data, rendering standard parameter estimation approaches unreliable.

### 2.2 Structural Identifiability Theory and Computational Analysis

Structural identifiability analysis provides the mathematical foundation for understanding when parameters can be uniquely determined from observed data. A parameter is *uniquely (or globally) structurally identifiable* if, given perfect observations (defined as noise-free and continuous for all time), there exists a unique parameter value that can produce such observations. A model is structurally identifiable if all parameters are individually globally identifiable [34–37].

Our analysis highlights a critical mathematical result [38]: while the standard SIR model (2.1) with observations of either active cases (*I*) or true incidence (*βSI/N*) maintains global structural identifiability for both *β* and *γ*, the extended system (2.1)—(2.2) becomes structurally unidentifiable under observations of detected incidence (*ϕβSI/N*). Specifically, *γ* remains uniquely identifiable, but *β* and *ϕ* cannot be separately determined—only their ratio *β/ϕ* is identifiable (see **Suppl. Materials S1**).

This identifiability crisis represents a fundamental computational limitation that affects a broad class of inverse problems in epidemiology and beyond, where scaling parameters confound the underlying system dynamics [39].

Despite the *β* and *ϕ* confounding, the effective reproduction number 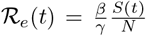 remains structurally identifiable from detected incidence (see **Suppl. Materials S1.3**).

### 2.3 Seroprevalence Integration as a Computational Solution

Seroprevalence data provide the mathematical constraint needed to resolve the identifiability crisis. Under ideal conditions (perfect sensitivity/specificity, representative sampling, persistent antibody detection), sero-prevalence directly measures the fraction of the population that has moved from susceptible to infected or recovered status, equivalent to knowing *S*(*t*) at survey time.

The computational power of this approach stems from complementary information: detected incidence captures temporal dynamics but suffers from unknown scaling (due to unknown *ϕ*), while seroprevalence provides absolute scale with limited temporal resolution. Together, they break the mathematical degeneracy that renders *β* and *ϕ* individually unidentifiable.

### 2.4 Computational Framework for Uncertainty Quantification

Our computational approach addresses parameter uncertainty through two complementary methods:

**Multi-start optimization** for noise-free scenarios identifies the manifold of parameter combinations that fit observed data perfectly, revealing the extent of identifiability issues.

**Bayesian inference with MCMC** quantifies uncertainty in realistic noisy conditions, providing full posterior distributions that capture both data limitations and prior knowledge.

The quantities of interest (QoIs) that drive epidemiological decision-making include: (i) state variables *S*(*t*), *I*(*t*), *R*(*t*), true incidence *I*_new_(*t*), and detected incidence 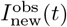 over time; (ii) case ascertainment ratio *ϕ*; (iii) basic reproduction number ℛ_0_ = *β/γ*; (iv) effective reproduction number ℛ_*e*_(*t*) = *β/γ S*(*t*)*/N*; (v) peak infections *I*_peak_ = max_*t*_ *I*(*t*) and timing *t*_peak_ = arg max_*t*_ *I*(*t*); (vi) final epidemic size *C*_∞_ = *N* − *S*_∞_.

## 3 Computational Results

### 3.1 Systematic Bias from Ignoring Case Underascertainment

We first establish the mathematical foundation underlying parameter estimation failures in epidemic models. When compartmental models are fitted to detected case data without accounting for incomplete surveillance—treating observed cases as complete truth—the resulting parameter estimates exhibit systematic bias that worsens as detection rates decline. This represents a model-data mismatch.

Figure 1 demonstrates this phenomenon computationally: fitting the SIR model to rescaled active case data (simulating various detection rates) produces visually acceptable fits while generating parameter errors exceeding 1000%. When only 1 in 50 cases is detected, the relative fitting error remains below 2%, yet relative errors in *γ, β*, and ℛ_0_ reach approximately 1000%, 500%, and 50%, respectively. This mathematical result extends to detected incidence data (**Supplementary Figure S1**), confirming that good fits can mask catastrophically wrong parameter estimates.

**Figure 1:**
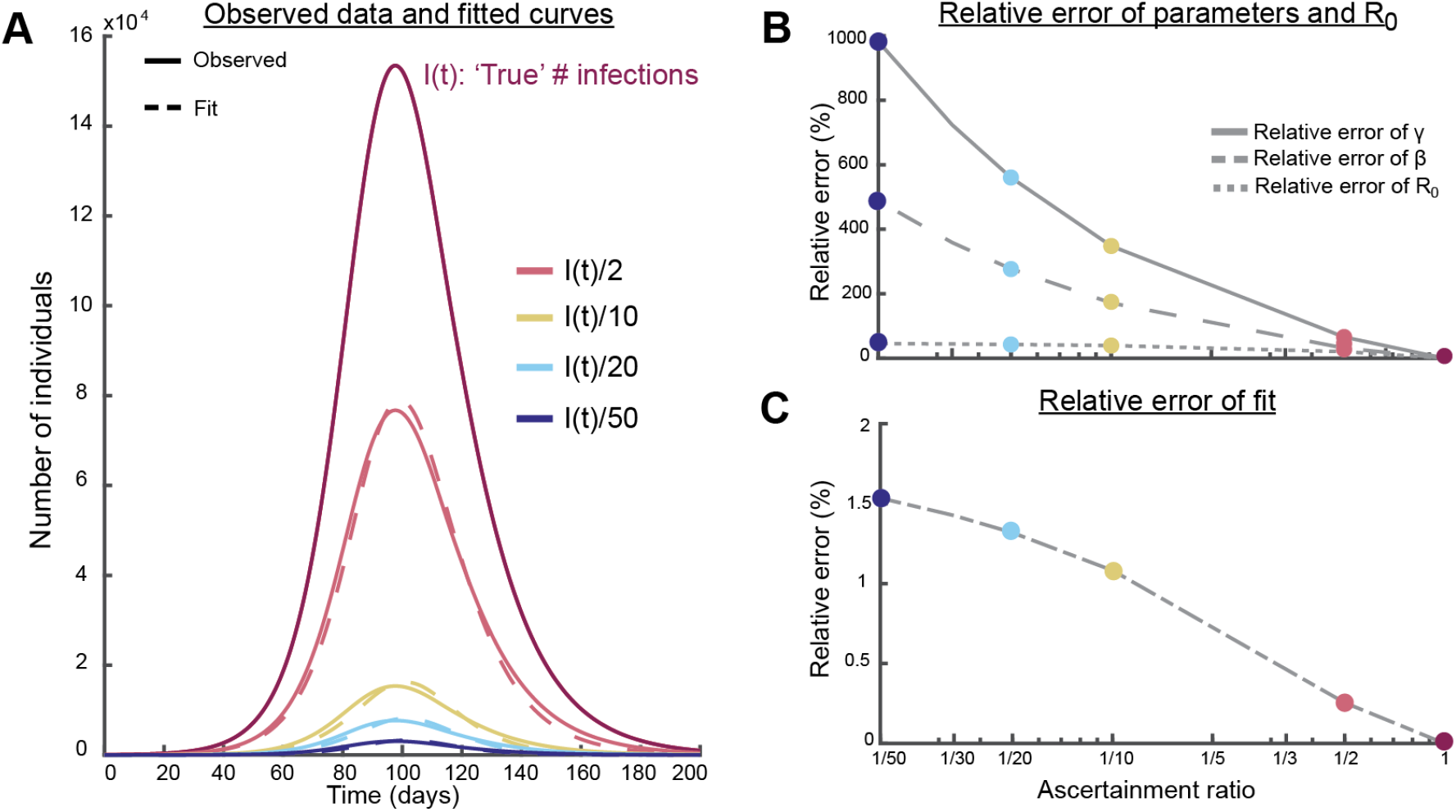
Computational demonstration that ignoring case underascertainment yields severe parameter bias despite good fits. **(A)** True infectious individuals *I*(*t*) (magenta) with rescaled curves (solid lines) and fitted estimates (dashed lines) for ascertainment ratios 1, 1/2, 1/20, and 1/50. **(B)** Relative parameter errors in *γ, β*, and ℛ_0_ as functions of ascertainment ratio. **(C)** Relative fitting errors for rescaled curves. Parameters: *β* = 0.2 day^−1^, *γ* = 0.1 day^−1^, (*S*(0), *I*(0), *R*(0)) = (*N* − 50, 50, 0), *N* = 10^6^.

### 3.2 Computational Implementation Framework

Our computational approach employs two complementary inference strategies tailored to different data scenarios and uncertainty quantification needs.

**Multi-start optimization** addresses the challenge of parameter identifiability in noise-free scenarios. We minimize the sum of squared errors between observations **Y** = [*y*_1_(*t*_1_), …, *y*_*n*_(*t*_*n*_)]^*T*^ and model predictions 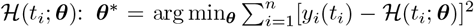. To comprehensively explore the parameter landscape, we perform over 1000 optimization runs with randomized initial conditions, accepting solutions that achieve visually perfect fits (SSE *<* 1000). This approach reveals the full manifold of parameter combinations consistent with observed data, providing computational proof of identifiability issues.

**Bayesian MCMC inference** quantifies uncertainty under realistic noisy conditions. We implement the posterior distribution *p*(***θ***|**Y**) ∝*p*(**Y**|***θ***)*p*(***θ***) using the No-U-Turn Sampler (NUTS), an adaptive Hamiltonian Monte Carlo variant that efficiently explores high-dimensional posteriors. The likelihood incorporates multiplicative Gaussian noise:

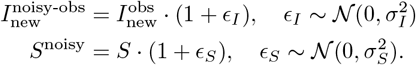

This computational framework enables systematic exploration of how data quality and quantity affect parameter identifiability, providing rigorous uncertainty quantification for epidemiological decision-making under realistic surveillance conditions.

### 3.3 Computational Characterization of Structural Unidentifiability

When models explicitly account for incomplete detection by inferring the ascertainment ratio *ϕ* alongside transmission parameters, they face the structural identifiability problem proven in Section 2. Our computational analysis reveals the practical consequences of this mathematical limitation.

**Figure 2B** shows that despite visually perfect fits to detected incidence data, the inferred trajectories of true incidence and compartment dynamics exhibit enormous variability. This occurs because the optimization algorithm finds multiple parameter combinations (*β, ϕ*) that produce identical detected incidence curves while implying vastly different epidemic dynamics. The model cannot distinguish among a continuous spectrum of scenarios ranging from high transmission rates with low detection ratios to low transmission rates with high detection ratios—all scenarios fit the data equally well.

**Figure 2:**
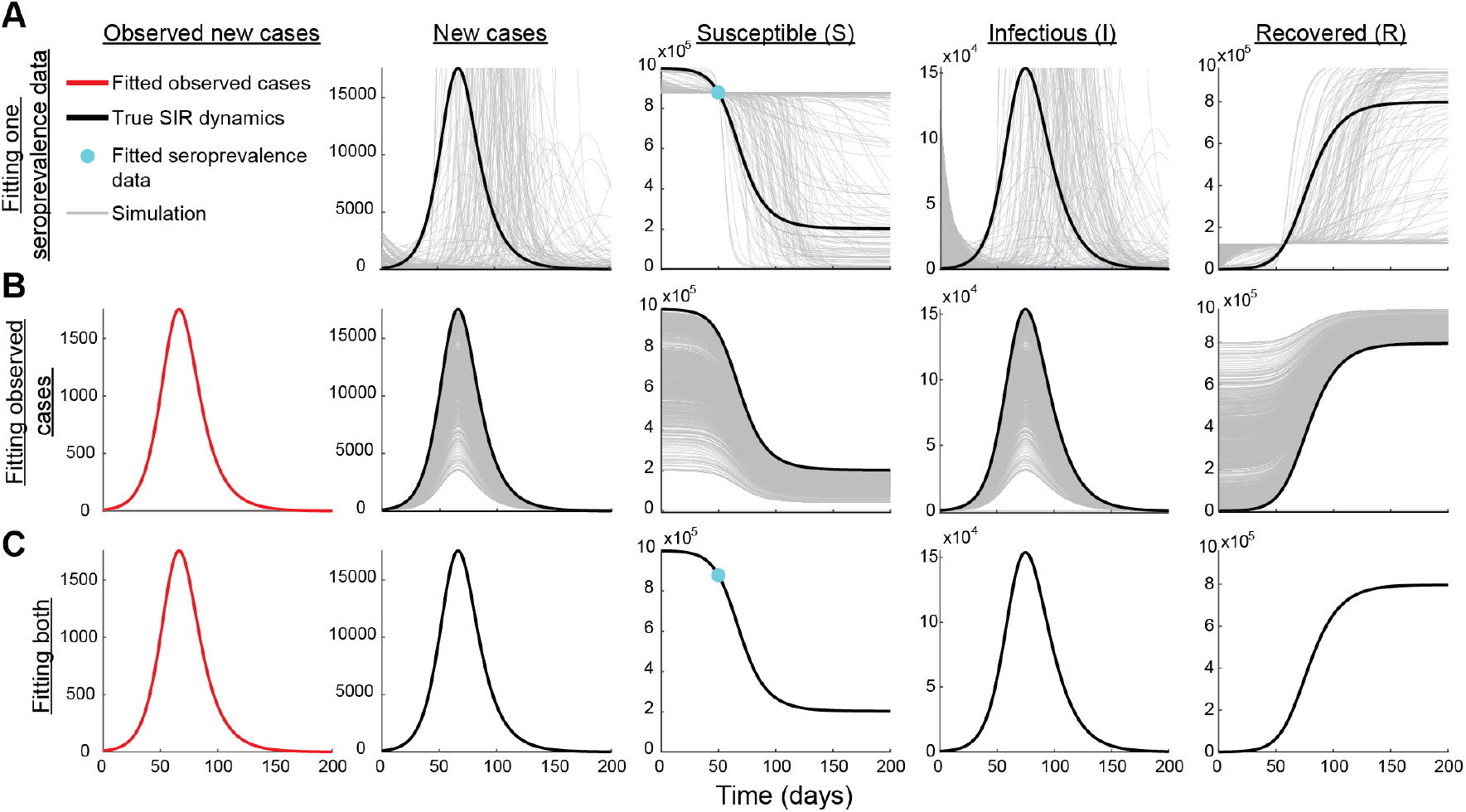
Computational analysis of identifiability: seroprevalence data resolves parameter degeneracy. Multi-start optimization results with parameters *β* = 0.2 day^−1^, *γ* = 0.1 day^−1^, (*S*(0), *I*(0), *R*(0)) = (*N* − *I*(0), 500, 0), *N* = 10^6^, *ϕ* = 0.1. **(A)** Single seroprevalence point (cyan dot) provides insufficient constraint, yielding wide variability in solution trajectories (gray lines) versus true dynamics (black curves). **(B)** Detected incidence alone (red curve) produces multiple equivalent solutions despite perfect fits. **(C)** Combined seroprevalence and detected incidence constrains solutions near true dynamics. We perform over 1,000 multi-start optimization runs with randomized initial conditions sampled within bounds *β, γ, ϕ* ∈ (0, 1) and *S*(0), *I*(0), *R*(0) ∈ [0, *N*] subject to *S*(0) + *I*(0) + *R*(0) = *N*.

This computational result confirms our theoretical identifiability analysis: *β* and *ϕ* are mathematically confounded, with only their ratio *β/ϕ* being uniquely determinable (**Supplementary Figure S2** and **Figure S3**). However, despite these underlying issues, fitted ℛ_*e*_(*t*) trajectories converge uniquely to true dynamics (**Supplementary Figure S4**), demonstrating its structural identifiability.

### 3.4 Algorithmic Solution Through Seroprevalence Integration

The mathematical resolution to identifiability comes through integrating seroprevalence measurements. **Figure 2C** demonstrates that even a single seroprevalence data point, alongside detected incidence, dramatically constrains parameter estimates, breaking the high correlation between *β* and *ϕ* (**Supplementary Figure S5**).

Our computational analysis reveals optimal data collection strategies: single seroprevalence measurements alongside detected incidence significantly improve inference, while three strategically timed seroprevalence measurements alone (early outbreak, epidemic peak, late stage) can reconstruct true dynamics with minimal uncertainty (**Supplementary Figure S6**).

To address concerns about the generalizability of our findings and ensure that the observed patterns are not artifacts of specific parameter choices, we conducted sensitivity analyses examining both parameter variations and measurement considerations. The qualitative features described in this section remain consistent across these variations. We show that the result of Figure 2C holds even when the ideal seroprevalence measurement is taken at different timepoints during an outbreak: incorporating even a single seroprevalence measurement dramatically reduces uncertainty in epidemiological quantities of interest regardless of when during the outbreak it is collected (**Supplementary Figure S7**). We assessed the impact of seroprevalence estimation errors and found that underestimating true seroprevalence by 10—30% leads to incorrect mapping from detected to true incidence, causing inferred quantities to deviate from their true values with increasing error in seroprevalence (**Supplementary Figure S8**). Varying transmission rates (corresponding to ℛ_0_: 2–5) and ascertainment ratio independently show that uncertainties in QoIs persist when only fitting to detected incidence (see **Supplementary Figure S9 and Figure S10**, respectively). Unlike Figure 2C, where trajectories generated from fitted parameters are systematically biased to one side of the true dynamics, varying the initial number of recovered individuals produces trajectories that fall on both sides of the true dynamics (**Supplementary Figure S11**), demonstrating that the observed bias in Figure 2C is an artifact of the ground truth data being generated under the assumption of no prior immunity in the population (i.e., *R*(0) = 0).

### 3.5 Bayesian Computational Framework for Noisy Data

Real epidemiological data contain measurement noise, requiring sophisticated uncertainty quantification methods. We implement Bayesian inference with MCMC to fully characterize parameter uncertainty under realistic conditions.

Figure 3 demonstrates that the identifiability solution remains robust under noise. Even with 40% noise in detected incidence and 10% noise in seroprevalence data, simultaneous fitting dramatically improves parameter estimation compared to using detected incidence alone.

Figure 4 provides a comprehensive quantification of uncertainty reduction across all epidemiological quantities of interest. The combined approach (orange violins) yields posterior distributions tightly concentrated around true values with dramatically reduced uncertainty compared to detected incidence alone (green violins), even at high noise levels. The distributions of the priors of the initial conditions and parameters for both noise-levels (10% and 40%) considered in this section, as well as their corresponding trace plots, are given in **Supplementary Section S11**.

The computational framework reveals that recovery rate estimates (*γ*) remain nearly identical regardless of data combination, consistent with our theoretical identifiability analysis. This parameter’s identifiability from temporal patterns makes it insensitive to seroprevalence addition. Similarly, the effective reproduction number (ℛ_*e*_(*t*)) maintains minimal uncertainty even from detected incidence alone (**Supplementary Figure S23**), consistent with its structural identifiability. In contrast, transmission-related parameters *β* and *ϕ* as well as initial conditions of the state variables show show dramatic improvement with seroprevalence integration.

**Figure 3:**
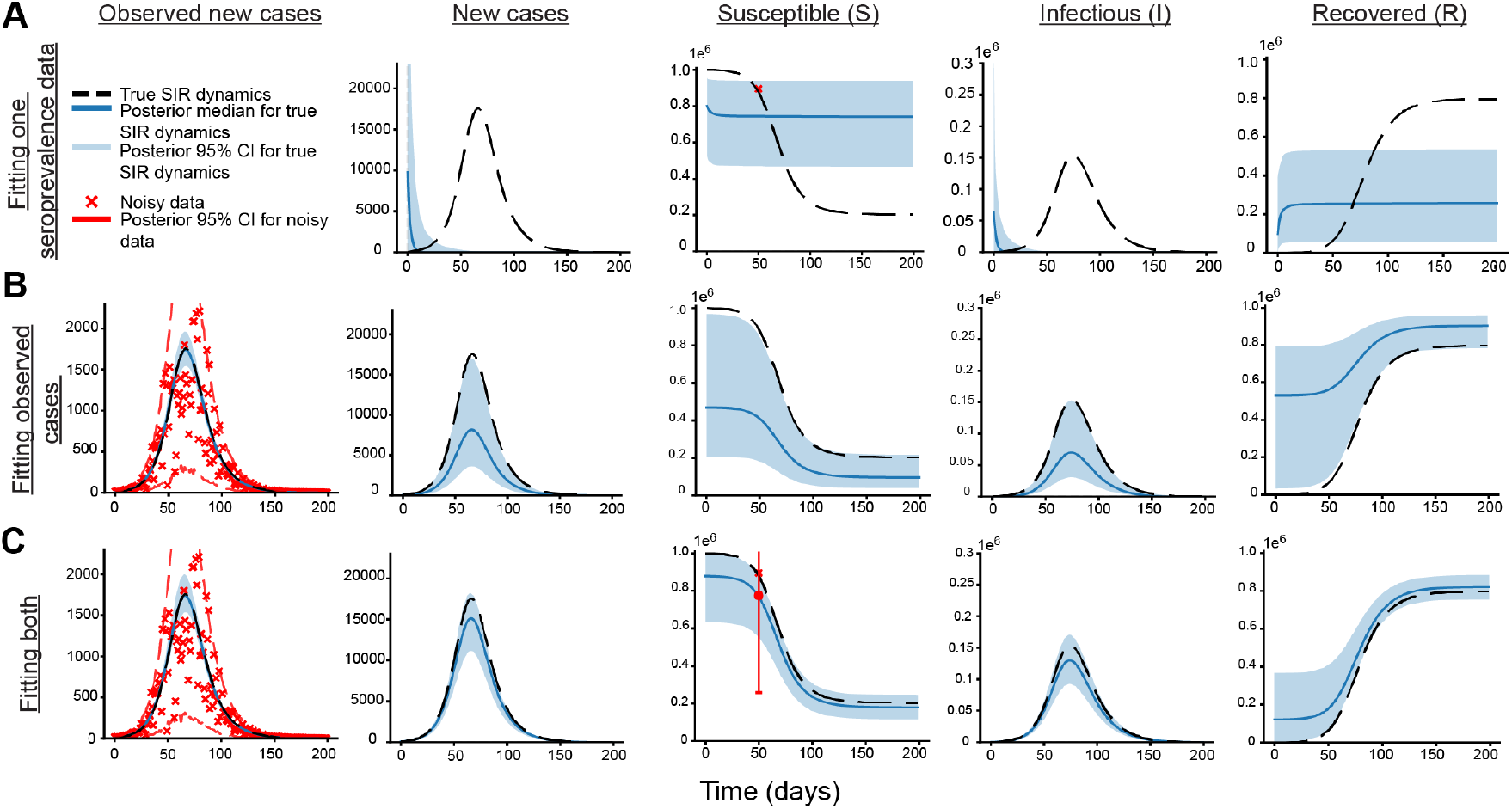
Bayesian computational analysis demonstrates robustness under realistic noise conditions. MCMC inference with *β* = 0.2 day^−1^, *γ* = 0.1 day^−1^, (*S*(0), *I*(0), *R*(0)) = (*N* − *I*(0) − *R*(0), 500, 0.1*N*), *N* = 10^6^, *ϕ* = 0.1. **(A)** Single noisy seroprevalence measurement yields wide 95% credible intervals (blue regions) deviating from true dynamics (dashed black). **(B)** Detected incidence with 40% noise (red crosses) produces wide uncertainty bands. **(C)** Combined noisy data substantially constrains posterior distributions.

**Figure 4:**
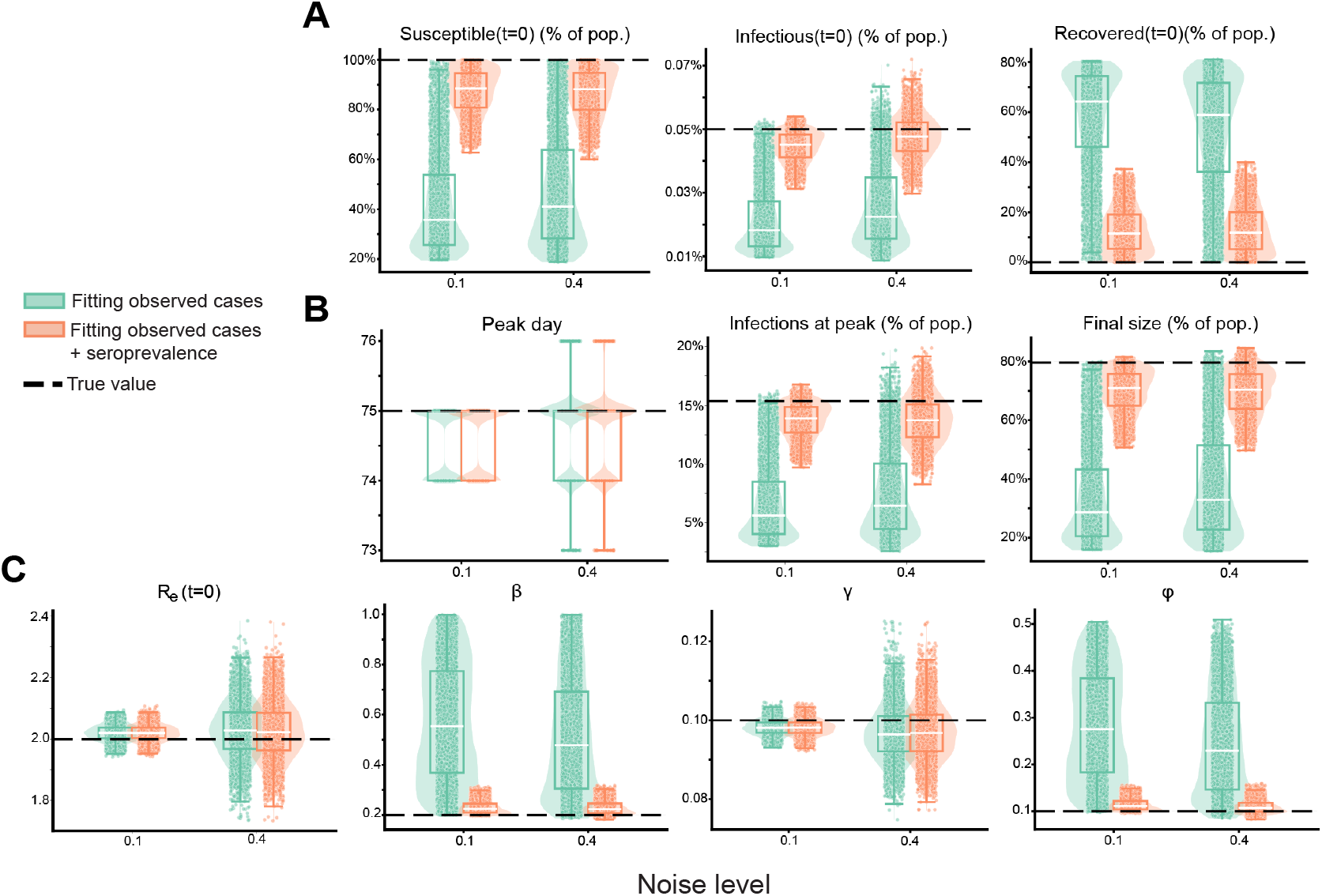
Comprehensive uncertainty quantification demonstrates systematic improvement across all epidemiological quantities. Posterior distributions comparing detected incidence alone (green violins: 10% noise; light green: included seroprevalence cases) versus combined data (orange violins: 40% incidence noise, 10% seroprevalence noise). Black dashed lines indicate true values. Seroprevalence integration consistently reduces uncertainty and improves accuracy across all QoIs. Parameters as in **Figure 3**.

## 4 Discussion

This study demonstrates that the integration of population-level seroprevalence data provides a mathematically rigorous and practically robust solution to the critical problem of parameter uncertainty in epidemiological models caused by case underascertainment. Our computational framework reveals two fundamental failures of models that rely solely on being fitted to detected case data. First, when models ignore underdetection, they produce catastrophically biased estimates of transmission rates and epidemic size, even when achieving deceptively reasonable fits to the observed data. Second, when models explicitly account for underdetection by including an ascertainment ratio as a parameter, they suffer from a structural unidentifiability crisis, where a wide spectrum of epidemiologically distinct scenarios can explain the data equally well. Our central finding is that incorporating even a single, well-timed seroprevalence measurement resolves both of these issues by providing an independent constraint on the cumulative scale of the epidemic, thereby breaking the mathematical degeneracy between the transmission rate and the ascertainment ratio.

An important nuance in our findings concerns the effective reproduction number, a critical metric for realtime outbreak assessment. Unlike the transmission rate and detection ratio, the effective reproduction number is identifiable from detected incidence alone. This has significant implications for the adequacy-for-purpose view of modeling, which evaluates models based on their fitness for specific intended uses rather than general accuracy [40–42]. A model fitted to detected cases may be adequate-for-purpose when the goal is determining whether an outbreak is growing or declining—the primary question guiding intervention timing. The effective reproduction number’s insensitivity to case ascertainment means that detected incidence data, despite being incomplete, can reliably inform this specific decision. However, the same model-data pairing becomes inadequate-for-purpose when inferential goals shift to estimating true infection rates, cumulative attack rates, or final epidemic size—quantities that directly depend on the unidentifiable individual parameters. This underscores how the adequacy of a data source must always be judged relative to the specific inferential task at hand.

The implications of these findings for public health and pandemic preparedness are substantial. Accurate estimation of infection rates, and epidemic size is critical because these quantities not only guide intervention strategies, vaccination targets, and resource allocation, but also shape our understanding of how diseases spread, evolve, and compare across pathogens and strains. We have computationally confirmed that relying on models fitted exclusively to reported case counts is an inherently flawed strategy that can lead to severely misguided policy decisions. By exposing this systematic vulnerability, our work underscores the necessity of moving beyond a single source of surveillance data. While several modeling studies include other data sources such as hospitalizations, deaths, or wastewater-based viral counts [25, 43, 44], these are also only proxies with uncertain scaling to true infections. In contrast, serology uniquely overcomes this limitation by directly measuring cumulative exposure. The integration of serological data transforms outbreak modeling from a potentially misleading exercise into a reliable tool for quantifying critical metrics such as the true infection rate, the effective reproduction number, and the final size of an epidemic. This enhanced accuracy is essential for effective resource allocation, intervention planning, and building resilient public health systems capable of responding to future pandemic threats with confidence.

The challenge of parameter unidentifiability, while highlighted here in an epidemiological context, represents a broader class of inverse problems prevalent across computational science [36, 37, 45–47]. In any system where the true underlying dynamics are observed through a process that involves unknown scaling factors or biases, standard inference methods can face substantial challenges. Our proposed solution—constraining the system with a complementary data source that provides information on absolute scale rather than temporal dynamics—offers a generalizable strategy for resolving such parameter degeneracies [48]. The computational framework, which combines multi-start optimization to map the landscape of possible solutions under a noise-free setting and Bayesian MCMC to quantify uncertainty under realistic noise, provides a transferable methodology for diagnosing and solving similar identifiability crises.

Our work highlights how synthetic experiments provide a powerful framework for assessing uncertainties in parameters, state variables, and derived quantities, offering a comprehensive picture of which aspects of system dynamics can be reliably inferred. When analytical identifiability becomes intractable due to model complexity, fitting to noise-free data remains computationally feasible and can reveal practical limitations of the model-data pairing in estimation and reconstruction of quantities of interest. Moreover, by systematically introducing observational noise into synthetic data, we can bridge the gap between ideal noiseless conditions and real-world complexity, isolating the impact of measurement error from structural model limitations. Real-world applications also introduce additional challenges through model-data mismatch, where simplifying model assumptions may fail to capture true data-generating processes [49]. Understanding the model’s ability to infer dynamical system behavior under synthetic noise-free and noisy conditions thus provides an essential foundation for interpreting discrepancies with real data and guiding model refinement, and should be undertaken as a first step for any new model as a matter of best practice, prior to fitting with real-world data.

A key strength of our findings is the demonstrated robustness of the solution under reasonable values of errors present in current surveillance systems. Our Bayesian inference framework showed that the dramatic reduction in parameter uncertainty holds even in the presence of substantial measurement noise, including up to 40% noise in detected incidence data and 10% in seroprevalence estimates. Furthermore, our sensitivity analyses showed that the benefits of seroprevalence integration are consistent across different transmission scenarios and are not highly dependent on the precise timing of the serological survey, strengthening the case for its broad implementation in routine surveillance. Both of these findings demonstrate that our approach is not merely a theoretical curiosity but a practically viable strategy for real-world application.

While this framework provides a clear path forward, we acknowledge several limitations that suggest avenues for future research. Our analysis relies on a simplified SIR model with a constant case ascertainment ratio, whereas real-world detection efforts and testing capacity fluctuate over time. Future work should extend this framework to models that incorporate such complexities. We also assume idealized seroprevalence measurements; further investigation is needed to quantify the impact of real-world complexities such as sampling biases, antibody waning [50–52], and imperfect test sensitivity and specificity [53–56]. Moreoever, while we demonstrate the powerful constraining effect of idealized serosurveys, the optimal design of real seroprevalence surveys, including determining the ideal timing, frequency, and sample size to maximize information gain, remains an important open question for future computational and epidemiological research. Finally, a limitation of our approach is that we have established that we can successfully reconstruct the dynamics (and infer the associated epidemiological parameters) of a fully observed disease outbreak. Future work should explore the extent to which population-level serological data can improve parameter inference in real-time, with access to incomplete data, such as the early stages of an outbreak and before the peak has been observed.

Our work provides not only a mathematical and computational solution to a long-standing problem in outbreak modeling but also a compelling economic and practical argument for implementing serosurveys in disease surveillance. With as few as 1,000 samples, reliable seroprevalence estimates can be obtained for populations of 1 million, at a total cost of about $69,000 ($69 per test [57]), provided true prevalence exceeds 2% [32, 58]. By enabling more accurate characterization of outbreak dynamics, this targeted data collection can prevent costly interventions and reduce the burden on healthcare systems, yielding a return on investment of several orders of magnitude while also informing policies that help safeguard lives. Therefore, integrating serological surveillance into routine epidemiological modeling should be viewed as both a mathematical necessity and a strategic investment in robust, evidence-based pandemic preparedness.

## 5 Methods

### 5.1 Synthetic Data Generation Framework

We generate controlled synthetic datasets using the SIR model (2.1) to evaluate our computational framework under known ground truth conditions. This approach enables rigorous assessment of identifiability issues and uncertainty quantification methods without the confounding effects of model misspecification or unknown data-generating processes.

#### 5.1.1 Noise-free synthetic data

Synthetic epidemic trajectories are generated by solving the SIR system (2.1) using MATLAB’s ode45 solver, an explicit Runge-Kutta (4,5) implementation based on the Dormand-Prince method.

Detected case data are generated by rescaling true trajectories according to the case ascertainment ratio *ϕ*. For detected active cases, we compute *I*^obs^(*t*) = *ϕ* × *I*(*t*), with initial conditions adjusted accordingly: *I*^obs^(0) = *ϕ* × *I*(0) and *S*(0) = *N* − (*I*^obs^(0) + *R*(0)), with *R*(0) = 0. For detected incidence experiments, the ascertainment ratio is incorporated through equation (2.2), with initial infectious population inferred during parameter estimation.

#### 5.1.2 Noisy synthetic data

To assess robustness under realistic surveillance conditions, we add multiplicative Gaussian noise to both detected incidence and seroprevalence data:

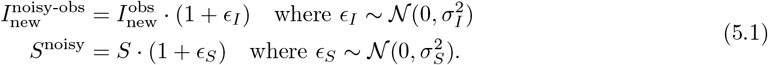

The noise parameters *σ*_*I*_ and *σ*_*S*_ represent multiplicative standard deviations. For example, *σ*_*I*_ = 0.1 corresponding to 10% noise where approximately 68% of observations fall within ±10% of true values. We refer to *σ*_*X*_ with *X* = {*I, S*} as *σ*% “percent noise” on *X*.

We set seroprevalence noise at 10% to reflect typical uncertainties in COVID-19 serological assays, where sensitivity ranges 85-95% and specificity ranges 95-99% [53, 54]. For detected incidence, we explore low noise (10%) and moderate noise (40%) scenarios, representing variability from test performance, data aggregation artifacts, and surveillance system inconsistencies—distinct from the systematic underascertainment modeled through *ϕ*.

### 5.2 Computational Inference Methods

#### 5.2.1 Multi-start optimization for noise-free analysis

Our multi-start optimization approach systematically explores parameter spaces to identify identifiability issues. We minimize the sum of squared errors between observations **Y** = [*y*_1_(*t*_1_), *y*_2_(*t*_2_), …, *y*_*n*_(*t*_*n*_)]^*T*^ and model predictions *ℋ* (*t*_*i*_; ***θ***):

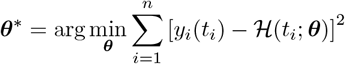

Implementation uses MATLAB’s `fminsearch, lsqcurvefit,` and `fmincon` optimizers. To comprehensively map the solution space, we perform over 1,000 optimization runs with randomized initial parameter guesses. Solutions achieving SSE *<* 1000 (corresponding to visually perfect fits) are retained for analysis. This approach reveals the complete manifold of parameter combinations consistent with observed data, providing computational evidence for theoretical identifiability results.

We highlight that in the case of fitting detected active case (Figure 1) or detected incidence data (Figure S1) to the SIR model without accounting for the reality that not all cases are observed, we use a multi-start approach to find parameter sets that minimize SSE the most.

In the context of multiple optimization runs with various initial guesses, the resulting distribution of parameter estimates does not represent true statistical uncertainty. This distribution merely reflects the sensitivity to initial guesses and the optimization algorithm’s convergence patterns rather than actual parameter uncertainty in the data. As a result, we do not display the histogram distribution of parameters using this method, nor do we use this method for the noise-present data fitting exercise (where a Bayesian method will be used).

#### 5.2.2 Bayesian inference for uncertainty quantification

For realistic noisy conditions, we implement Bayesian inference to quantify parameter uncertainty. The posterior distribution *p*(***θ*** |**Y**) ∝*p*(**Y** |***θ***)*p*(***θ***) combines the likelihood defined by the noise model (5.1) with uniform priors on parameters and initial conditions.

Prior specifications include *β, γ, ϕ* ~𝒰 ([0.01, 1]) for model parameters and *S*(0), *I*(0), *R*(0) ~𝒰 (0, *N*) subject to *S*(0) + *I*(0) + *R*(0) = *N*. Initial condition priors are implemented using Dirichlet distributions with concentration parameters *α* = [1, 1, 1] then rescaled by *N*, providing equivalent uniform constraints with improved computational efficiency.

Noise parameters *σ*_*I*_, *σ*_*S*_ ~𝒰 ([0.001, 0.5]) are jointly estimated, with full posterior distributions provided in Supplementary Materials.

#### 5.2.3 MCMC implementation

Sampling uses the No-U-Turn Sampler (NUTS), an adaptive Hamiltonian Monte Carlo variant implemented in NumPyro [59]. Our sampling protocol includes:

- Preliminary warmup: 2,000 iterations (step size 1e-3, target acceptance 0.4)
- Burn-in: 5,000 iterations (discarded)
- Final warmup: 2,000 iterations (step size 1.0, target acceptance 0.9)
- Posterior sampling: 5,000 iterations for analysis

This custom implementation ensures robust convergence in scenarios with complex identifiability structures. Simpler default schemes suffice when both seroprevalence and case data are available, but the enhanced protocol handles challenging unidentifiable cases. Convergence assessment uses visual trace plot inspection, with full diagnostics in Supplementary Materials.

### 5.3 Structural Identifiability Analysis

Structural identifiability analysis uses two complementary symbolic computation tools: DAISY [60] and Julia’s `StructuralIdentifiability.jl` [61]. We use these tools to determine parameter identifiability as theoretical properties independent of data quality or estimation algorithms, while assuming unknown initial conditions during structural identifiability analysis.

### 5.4 Parameter Ranges and Computational Settings

All experiments use population size *N* = 10^6^ individuals. Baseline parameters are *β* = 0.2 day^−1^, *γ* = 0.1 day^−1^ (corresponding to ℛ_0_ = 2), with case ascertainment ratio of *ϕ* = 0.1. Sensitivity analyses explore transmission rates *β* ∈ [0.2, 0.5] day^−1^ (ℛ_0_ ∈ [2, 5]) with case ascertainment ratios ranging from *ϕ* = 0.1 to *ϕ* = 1. Initial conditions and parameters vary by experiment, with specific values provided in figure captions. The computational framework is designed to handle diverse parameter regimes while maintaining theoretical rigor and practical applicability.

## Data Availability

All data produced in the present study are available upon reasonable request to the authors

## Acknowledgements

Funding for this work was made possible in part by cooperative agreement CDC-RFA-FT-23-0069 from the CDC’s Center for Forecasting and Outbreak Analytics. The views expressed in this publication do not necessarily reflect the official policies of the Department of Health and Human Services/Centers for Disease Control and Prevention. AN was supported by the CDC-CFA award for the Atlantic Coast Center for Infectious Disease Dynamics and Analytics (ACCIDDA): NU38FT000012-01-00 (UNC). MEL was supported by the Eric and Wendy Schmidt Center at the Broad Institute of MIT and Harvard and Basis Research Institute.

