## Supplementary material for "Resolving Parameter Uncertainty in Outbreak Models Through Population-Level Serological Surveillance": supp

**S2 Fitting to Observed Active Cases without Accounting for Case Detection ratio** **6**

**S3 Fitting to Detected Incidence without Accounting for Case Detection Ratio** **6**

**S4 Fitting to Noise-free Data** **7**

**S5 Fitting up to Three Seroprevalence Data Only** **11**

**S6 Fitting Detected Incidence and Single Seroprevalence: Sensitivity to Seroprevalence Timing** **12**

**S7 Underestimating Seroprevalance** **13**

**S8 Varying Transmission Rate Only** **14**

**S9 Varying Case Ascertainment Ratio Only** **15**

**S10Varying Initial Number of Recovered Individuals** **16**

**S11 Bayesian Inference on Noisy Data** **17**

### S1 Structural Identifiability

Structural identifiability analysis is carried out using two software packages, namely DAISY [1] and Julia `StructuralIdentifiability.jl` [2]. We refer readers to Chowell et al. [3] and Liyanage et al. [4] for primers on using DAISY and Julia for epidemic models, respectively.

#### S1.1 Identifiability of SIR model with respect to detected incidence

The SIR model under the observation of the detected incidence (i.e., model (2.1)-(2.2)) is structurally unidentifiable. We present the methodology briefly below (see [3] for further details).

We run the following DAISY code on Version 2.1, where we define `y` as the observation (here, detected incidence $\phi\beta SI/N$):

```
let N = 1$ % we assume N is known

B_:={Y,S,I,R}$ % variable vector
B1_:={beta,gamma,phi}$ % unknown parameter vector

NX_:=3$ % number of states
NU_:=0$ % number of inputs
NY_:=1$ % number of output

%model equations

C_:= {df(S,t) = - beta*S*I/N,
      df(I,t) = beta*S*I/N- gamma*I,
      df(R,t) = gamma*I,
            y = phi*beta*S*I/N}$
FLAG_:=1$
daisy()$

ICK_:={S=1,I=2,R=3}$  %IC
ICUNK_:={}$
CONDINIZ()$
END$
```

DAISY will show that the model is nonidentifiable when initial conditions are not known. To investigate the relationship between parameters, we proceed as follows.

Using DAISY, we get the following input-output equation:

```
df(y,t,2)**2*y**2*phi**2 - 2*df(y,t,2)*df(y,t)**2*y*phi**2
+df(y,t,2)*df(y,t)*y**2*gamma*phi**2 + 4*df(y,t,2)*y**4*beta*phi
+ df(y,t,2)*y**3*gamma**2*phi**2 + df(y,t)**4*phi**2 - df(y,t)**3*y*gamma*phi**2
- 4*df(y,t)**2*y**3*beta*phi - df(y,t)**2*y**2*gamma**2*phi**2
+ 2*df(y,t)*y**4*beta*gamma*phi + 4*y**6*beta**2 +y**5*beta*gamma**2*phi$
```

Dividing the right-hand side by $\phi^2$ gives the following monic polynomial:

$$(y'')^2y^2 - 2y''(y')^2y + 4y''y'y^2\gamma + (y'')^3\gamma^2 + (y')^4 - (y')^3y\gamma + 2y'y^4\beta\gamma/\phi + 4y^6\beta^2/\phi^2 + y^5\beta\gamma^2/\phi = 0 \quad \text{(S1.1)}$$

If the model is not structurally identifiable, then there exist two parameter sets $(\beta_1, \gamma_1, \phi_1)$ and $(\beta_2, \gamma_2, \phi_2)$

that produce identical monic polynomial (S1.1). Comparing the like terms of the monic polynomial associated with these parameter sets gives:

$$\gamma_1 = \gamma_2 \quad \text{and} \quad \beta_1/\phi_1 = \beta_2/\phi_2.$$

Consequently, the parameter $\gamma$ is globally identifiable even when the initial conditions of the SIR model are unknown. Meanwhile, the parameters $\beta$ and $\phi$ are unidentifiable; however, their ratio $\beta/\phi$ is globally identifiable.

#### S1.2 Identifiability of SIR model with respect to active cases, incidence, and seroprevalence

The SIR model (2.1) is uniquely structurally identifiable under three observation scenarios: active cases ($I$), prevalence ($\beta SI/N$), or ideal seroprevalence (which we have defined to as equivalent to observing $S$). For each scenario, we modify y in the DAISY code from Section S1.1 respectively as:

```
y = I
```

```
y = beta S*I/N
```

```
y = S
```

In each of these scenerios, DAISY will show:

```
MODEL GLOBALLY IDENTIFIABLE$

INITIAL CONDITION(S) NOT NECESSARY$
```

For completeness and for consistency's sake, we provide the input-output equation obtained for observation being $I$, $\beta SI/N$, and $S$, as follows:

```
df(y,t,2)*y - df(y,t)**2 + df(y,t)*y**2*beta + y**3*beta*gamma
```

```
df(y,t,2)**2*y**2 - 2*df(y,t,2)*df(y,t)**2*y + df(y,t,2)*df(y,t)*y**2*gamma
+ 4*df(y,t,2)*y**4*beta + df(y,t,2)*y**3*gamma**2 + df(y,t)**4 - df(y,t)**3*y*gamma
- 4*df(y,t)**2*y**3*beta - df(y,t)**2*y**2*gamma**2 + 2*df(y,t)*y**4*beta*gamma
+ 4*y**6*beta**2 + y**5*beta*gamma**2
```

```
- df(y,t,2)*y + df(y,t)**2 + df(y,t)*y**2*beta - df(y,t)*y*gamma
```

The monic polynomials corresponding to about input-output equation are respectively given by:

$$y''y - (y')^2 + \beta y'y^2 + \beta\gamma y^3 = 0,$$
$$(y'')^2y^2 - 2y''(y')^2y + \gamma y''y'(y)^2 + 4\beta y''y^4 + \gamma^2(y'')y^3 + (y')^4 - \gamma(y')^3y - 4\beta(y')^2y^3 - \gamma^2(y')^2y^2 + 2\beta\gamma y'y^4 + 4\beta^2y^6 + \beta\gamma^2y^5 = 0,$$
$$y''y - (y')^2 - \beta y'y^2 - \gamma y'y = 0.$$

Using similar argument as Section S1.1, for each observation we have

$$\beta_1 = \beta_2 \quad \text{and} \quad \gamma_1 = \gamma_2.$$

#### S1.3 Identifiability of the effective reproduction number

The effective reproduction number, $\mathcal{R}_e(t)$, of the SIR model is given by:

$$\mathcal{R}_e(t) = \frac{\beta}{\gamma}\frac{S(t)}{N}.$$

Using Julia's , we show that the effective reproduction number is uniquely identifiable under the observation of detected incidence as follows. Since we assume the total population $N$ is known, without loss of generality, let $N = 1$. We have the following model in Julia.

We use Julia's `StructuralIdentifiability.jl` package [2] to analyze the structural identifiability of $\mathcal{R}_e(t)$ under the observation of detected incidence $\phi\beta S(t)I(t)/N$. Since we assume the total population $N$ to be known, without loss of generality, let $N = 1$. The model is implemented in Julia as follows:

```
ode = @ODEmodel(
x1'(t) = -beta*x1(t)*x2(t)/(1),
x2'(t) = beta*x1(t)*x2(t)/(1) - gamma*x2(t),
x3'(t) = gamma*x2(t),
y(t) = phi*beta*x1(t)*x2(t)/(1)
)
```

where x1(t), x2(t), and x3(t) correspond to $S(t), I(t)$, and $R(t)$, respectively, and y(t) represents the observed detected incidence.

To assess the structural identifiability of $\mathcal{R}_e(t)$, we use the following command:

```
assess_identifiability(ode, funcs_to_check = [beta//gamma * x1/1])
```

This yields:

```
OrderedCollections.OrderedDict{Any, Symbol} with 1 entry:
  (x1(t)*beta)//gamma => :globally
```

demonstrating that $\mathcal{R}_e(t)$ is globally structurally identifiable under the observation of detected incidence. The fact that $\mathcal{R}_e(t)$ trajectories obtained from fitting the model to noise-free detected incidence data are visually indistinguishable from the true $\mathcal{R}_e(t)$ (see Supplementary Figure S4) provides computational confirmation that $\mathcal{R}_e(t)$ is structurally identifiable under the observation of detected incidence, since identifiability under data implies structural identifiability.

### S2 Fitting to Observed Active Cases without Accounting for Case Detection ratio

**Table S1:** Parameter estimation results for different ascertainment ratios. The true values of $\beta$ and $\gamma$ are 0.2 day$^{-1}$ and 0.1 day$^{-1}$, respectively. We drop the units of $\beta$ and $\gamma$ in the table below for simplicity.

| Ascertainment Ratio | $\beta$ | $\gamma$ | $\mathcal{R}_0$ | Rel. Err. $\beta$ (%) | Rel. Err. $\gamma$ (%) | Rel. Err. $\mathcal{R}_0$ (%) | Fit Error (%) |
|---|---|---|---|---|---|---|---|
| 1 | 0.200 | 0.100 | 2.00 | 0.1 | 0 | 0 | 0 |
| 1/2 | 0.263 | 0.166 | 1.588 | 31.645 | 65.804 | 20.602 | 0.256 |
| 1/10 | 0.543 | 0.448 | 1.211 | 171.454 | 348.144 | 39.427 | 1.082 |
| 1/20 | 0.755 | 0.660 | 1.14 | 277.325 | 560.439 | 42.868 | 1.318 |
| 1/30 | 0.917 | 0.823 | 1.114 | 358.699 | 723.423 | 44.294 | 1.425 |
| 1/50 | 1.176 | 1.082 | 1.087 | 487.842 | 981.940 | 45.668 | 1.534 |

### S3 Fitting to Detected Incidence without Accounting for Case Detection Ratio

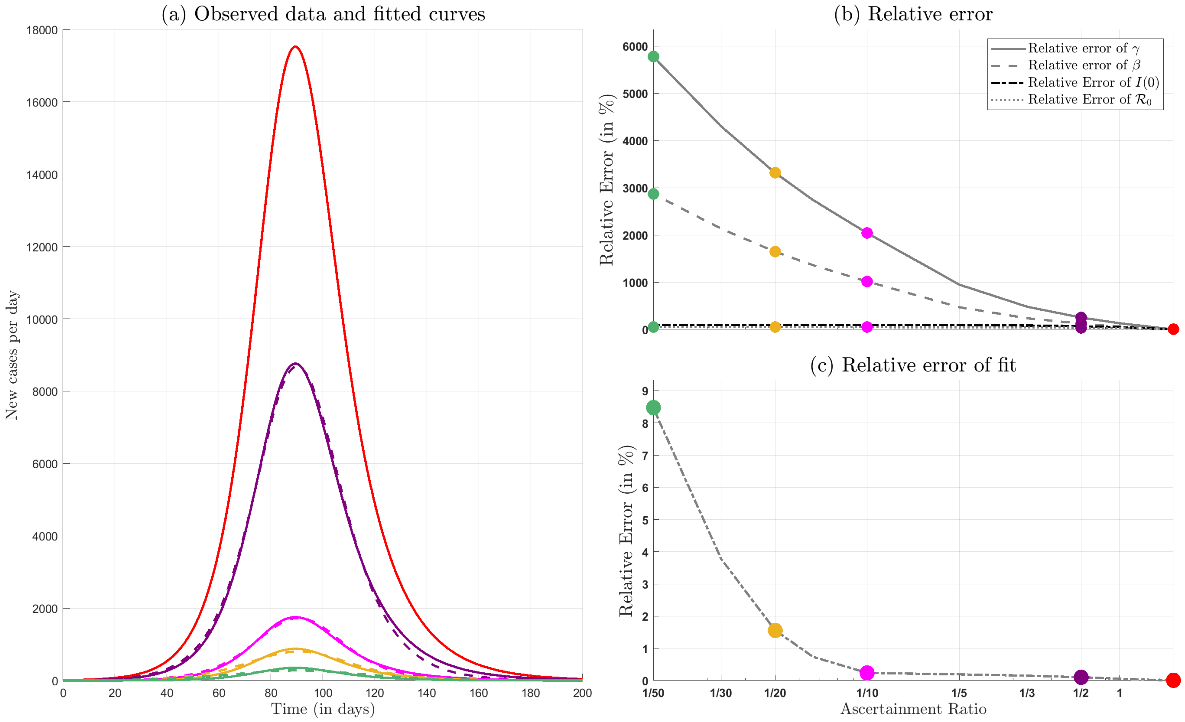

**Figure S1: Performing data-fitting without accounting for ascertainment (case detection) ratio can severely impact parameter inference, even when the fitted new case curve appears accurate due to low relative error. (a)** The number of true new cases, $I_{\text{new}}(t) = \beta SI/N$, generated from the SIR model along with the rescaled $I_{\text{new}}(t)$ curves (given with solid lines) and their respective fitted curves (given with dashed lines). **(b)** The curves showing relative error in recovery rate ($\gamma$), infection rate ($\beta$), initial number of infectious individuals ($I(0)$), and basic reproduction number ($\mathcal{R}_0$) for various values of case ascertainment ratio. **(c)** The relative error of the fit, in comparison to the synthetic data, for various values of the ascertainment ratio. Unlike Figure 1, we also choose to fit $I(0)$.

### S4 Fitting to Noise-free Data

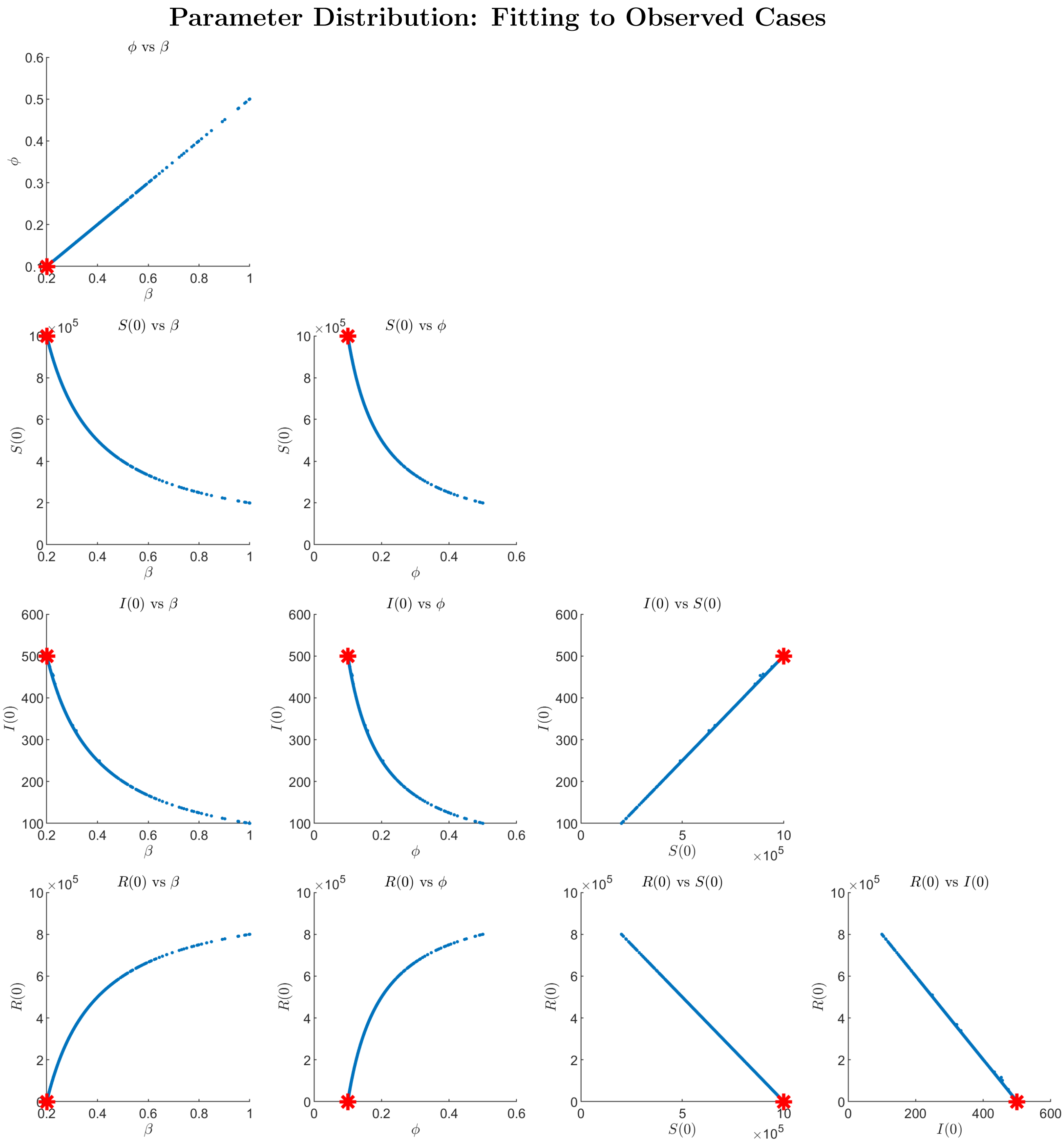

**Figure S2: Fitting an SIR model to detected/observed new case data without knowing the case detection ratio can lead to parameters being unidentifiable.** Each panel shows the relationship between two parameters, with red stars indicating true parameter values. The top-left panel shows the direct proportional relationship between $\phi$ and $\beta$, demonstrating non-identifiability. Other panels illustrate how initial conditions ($S(0)$, $I(0)$, $R(0)$) relate to these epidemic parameters and to each other, forming a connected manifold of equally good fits. This structural non-identifiability means multiple parameter combinations can produce identical fits to observed data, making accurate parameter estimation impossible without additional information about the case detection ratio.

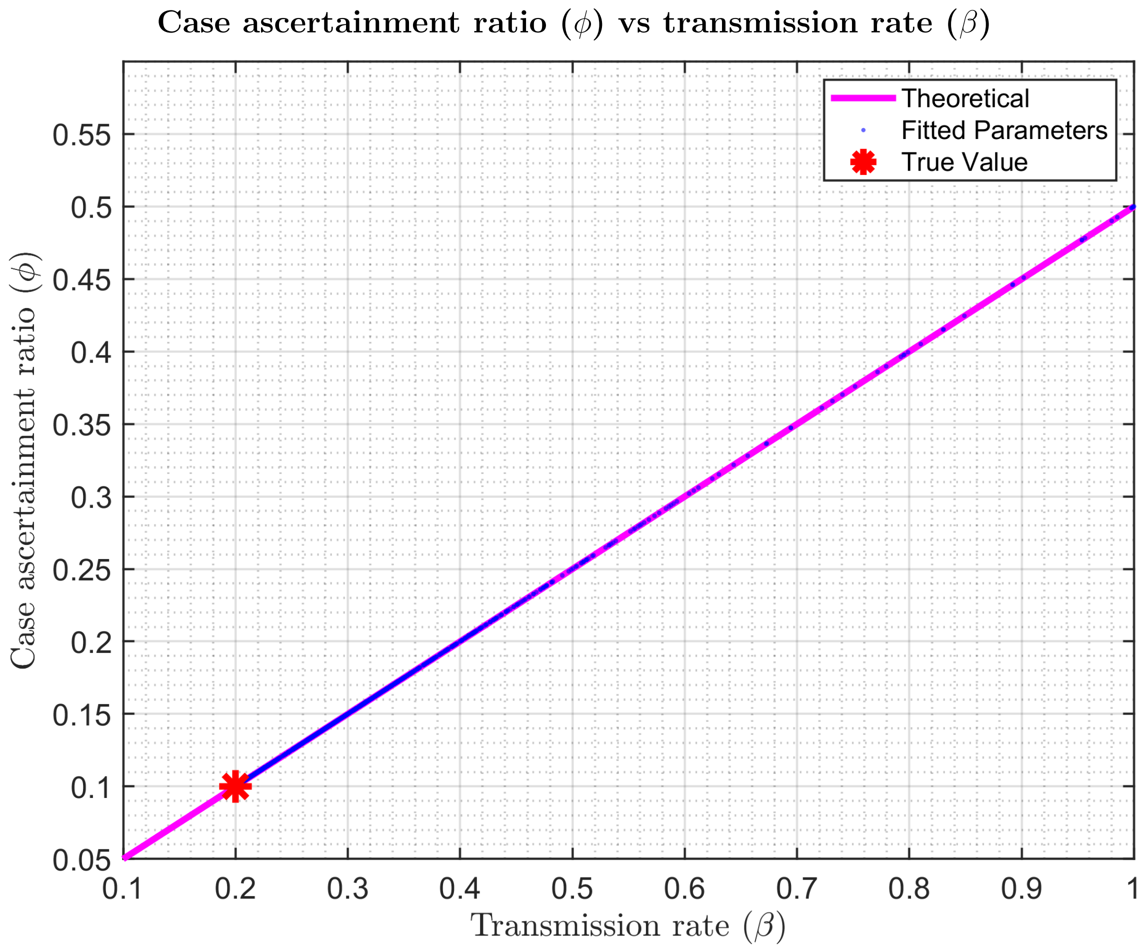

**Figure S3: Parameter identifiability analysis showing preservation of the $\beta/\phi$ ratio when SIR model is fitted to noise-free detected incidence data.** Estimated $\beta$ and $\phi$ values (blue dots) cluster tightly along the theoretical line $\beta/\phi$ (magenta), confirming that while individual parameters are not uniquely identifiable, their ratio is well-preserved. True values ($\beta = 0.2$ day$^{-1}$, $\phi = 0.1$) are shown as a red star. See Section S1.1) for further discussion on the theoretical result.

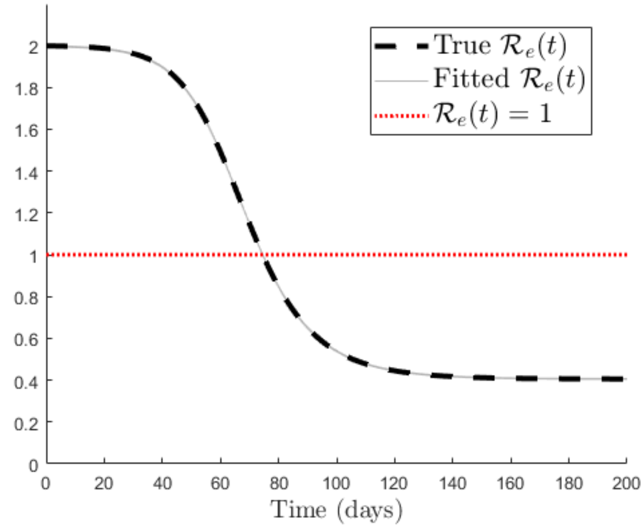

**Figure S4: Structural identifiability of $\mathcal{R}_e(t)$ confirmed through noise-free data fitting.** The effective reproduction number $\mathcal{R}_e(t)$ obtained from fitting the SIR model to noise-free detected incidence data (solid gray line) is visually indistinguishable from the true $\mathcal{R}_e(t)$ trajectory (dashed black line), providing computational confirmation of structural identifiability. The horizontal dotted red line marks the critical epidemic threshold $\mathcal{R}_e(t) = 1$, where values above 1 indicate epidemic growth and values below 1 indicate decline.

#### Parameter distribution: simultaneously fitting to observed case data and one seroprevalence data

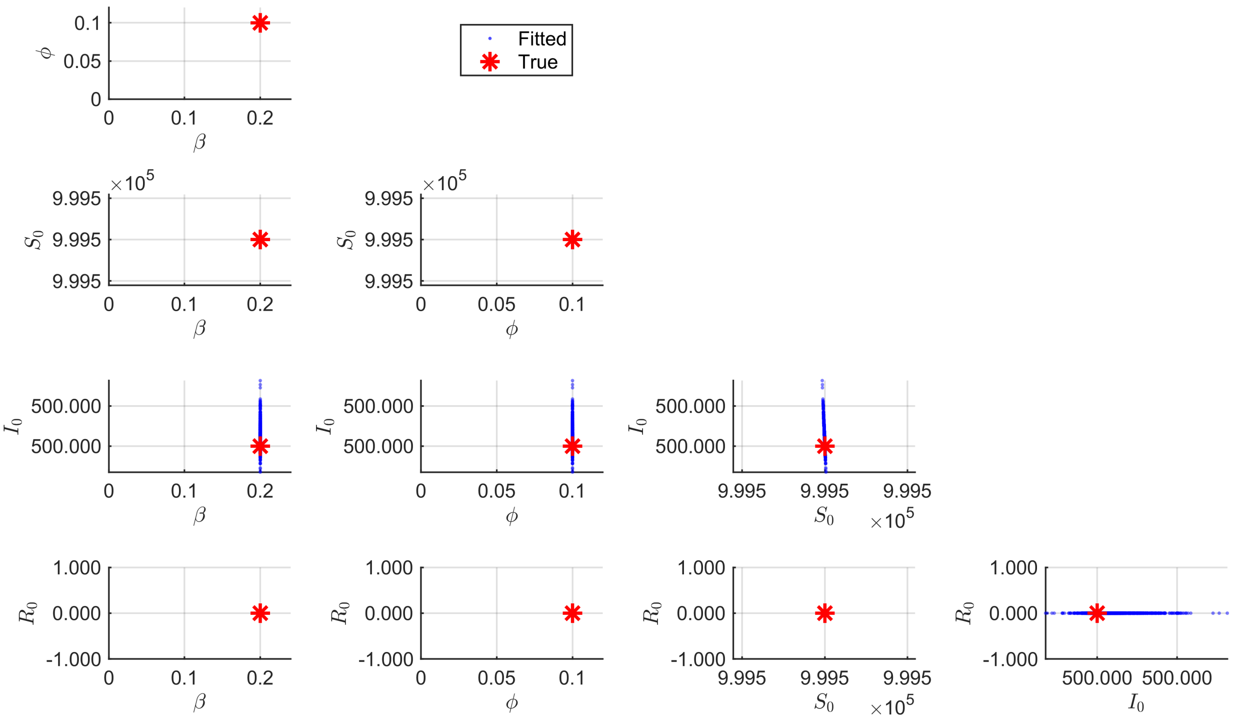

**Figure S5: Simultaneously fitting a mechanistic model to noise-free seroprevalance and detected case data dramatically improves characterization of epidemiological QoIs.** Fitting to one perfect population-level seroprevalence data, in addition to detected incidence, leads to perfect recovery of parameters and initial conditions. The blue dots represent individual parameter sets from accepted model fits, while the red stars indicate the true parameter values. The plots show the pairwise relationships between the transmission rate ($\beta$), ascertainment ratio ($\phi$), initial susceptible population ($S_0$), initial infected population ($I_0$), and initial recovered population ($R_0$). For simplicity, we assume the recovery rate ($\gamma$) is known.

### S5 Fitting up to Three Seroprevalence Data Only

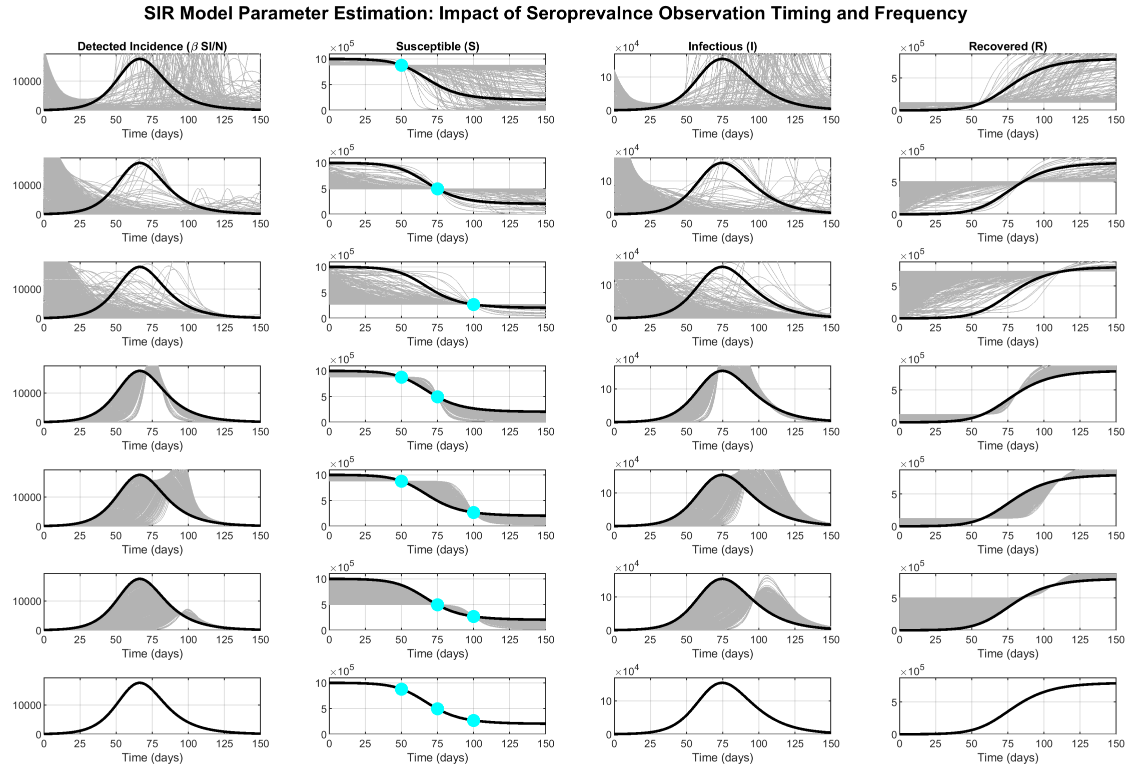

**Figure S6: Impact of seroprevalence observation timing and frequency on SIR parameter estimation uncertainty.** Each row represents a different observation scenario: single observations at day 50, 75, or 100 (rows 1-3); paired observations at days (50,75), (50,100), or (75,100) (rows 4-6); and triple observations at days (50,75,100) (row 7). Column 1 shows detected incidence ($\beta SI/N$), while columns 2-4 display the susceptible (S), infectious (I), and recovered (R) compartments. Cyan circles in column 2 represent seroprevalence observation points used for parameter fitting. Black curves represent the true SIR dynamics, while gray curves show all accepted parameter fits that satisfy the goodness-of-fit criteria. The true dynamics are generated with parameters $\beta = 0.2$ day$^{-1}$, $\gamma = 0.1$ day$^{-1}$, and initial conditions $(S(0), I(0), R(0)) = (N - I(0), 500, 0)$, where $N = 10^6$ individuals. Increasing the number and strategic timing of seroprevalence measurements substantially reduces parameter estimation uncertainty, with the greatest improvement achieved through observations spanning early outbreak, epidemic peak, and late-stage dynamics.

#### S6 Fitting Detected Incidence and Single Seroprevalence: Sensitivity to Seroprevalence Timing

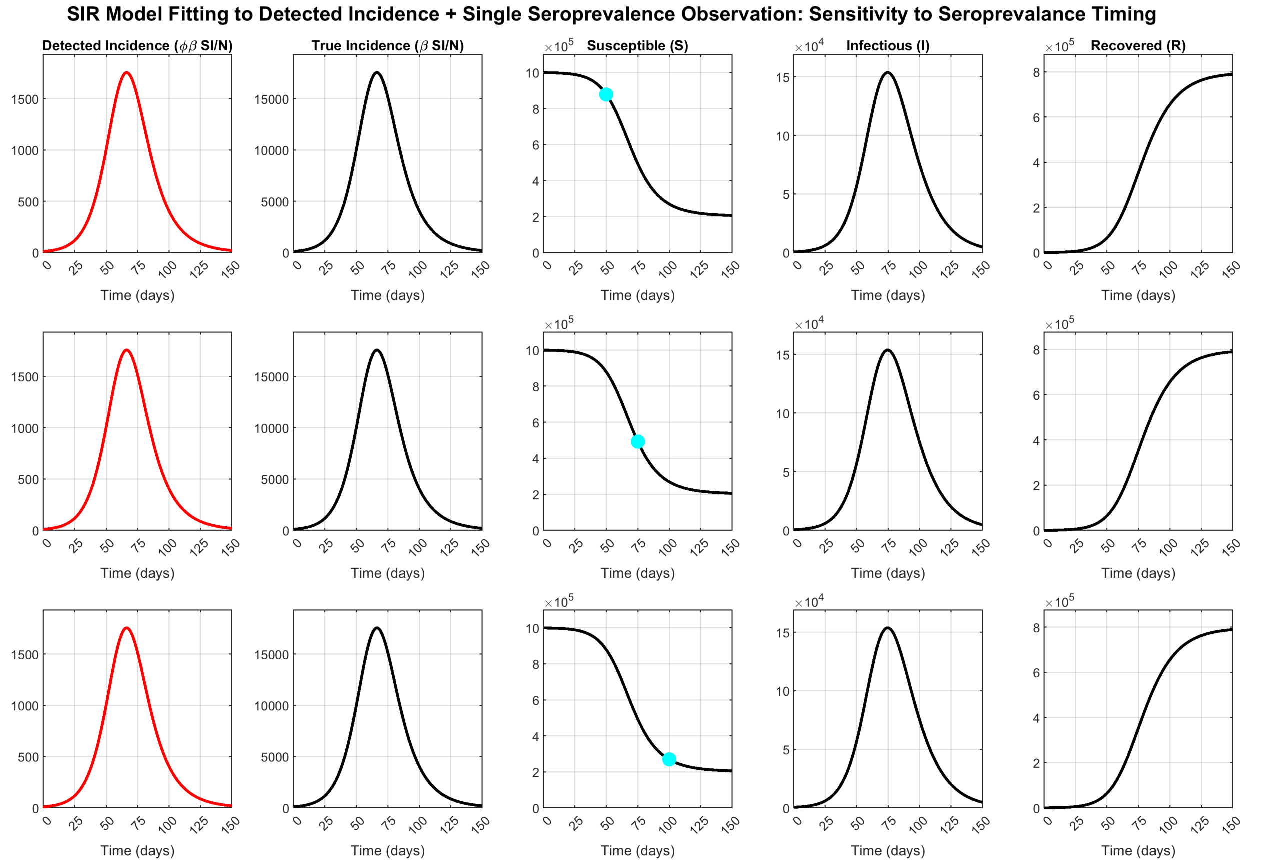

**Figure S7: Robustness of dual data fitting across different seroprevalence observation times.** The three rows demonstrate that the findings from Figure 2C hold consistently regardless of when the single seroprevalence observation is taken: at $t = 50$, $t = 75$, and $t = 100$ days. In all scenarios, fitting to both detected incidence curve ($\phi\beta SI/N$) and a single ideal seroprevalence measurement (cyan circles) successfully constrains parameter estimates and nearly reproduces the true dynamics across all quantities of interest. The gray curves represent all accepted parameter fits, while the true dynamics (black and red curves) are generated with parameters $\beta = 0.2$ day$^{-1}$, $\gamma = 0.1$ day$^{-1}$, $\phi = 0.1$, and initial conditions $(S(0), I(0), R(0)) = (N - I(0), 500, 0)$, where $N = 10^6$ individuals. The gray curves are underneath the red and black curves.

### S7 Underestimating Seroprevalance

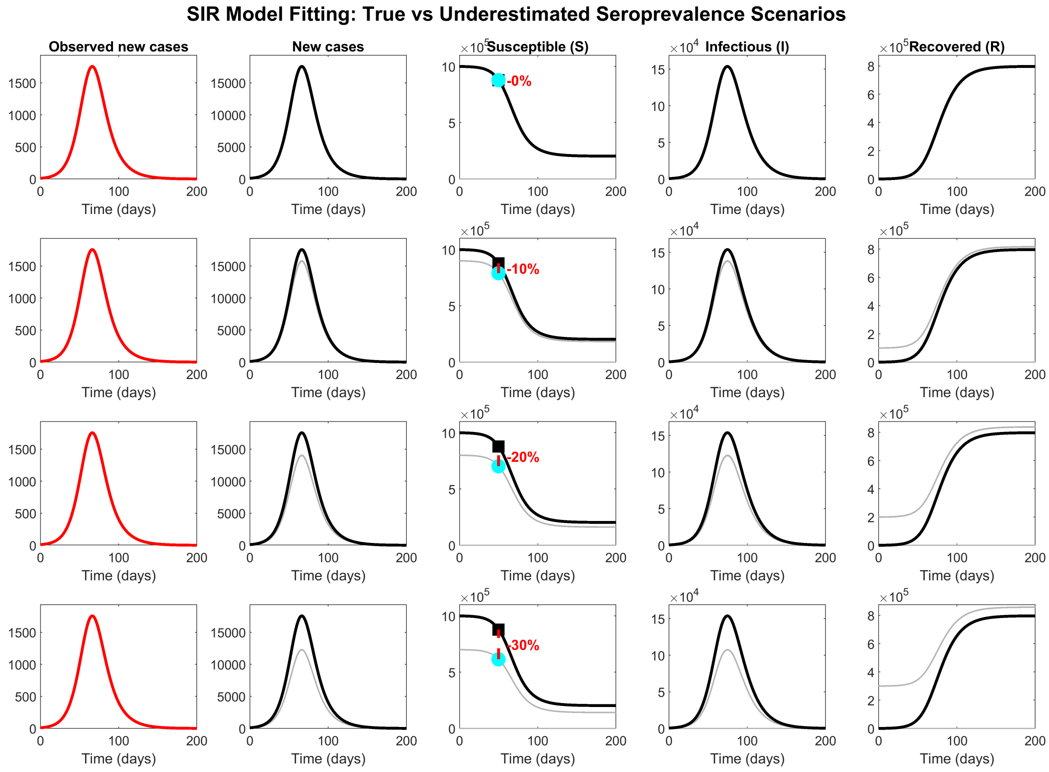

**Figure S8: Larger error in seroprevalance leads to larger error in QoIs.** The first row is a replication of Figure 2C, while the remaining mimick Figure 2C under the assumption of 10%, 20%, and 30% underestimation of population-level seroprevalance data. The curves obtained through data fitting are depicted with a shaded gray color. The true dynamics, given with black curves, are generated with parameters $\beta = 0.2$ day$^{-1}$, $\gamma = 0.1$ day$^{-1}$, and initial conditions $(S(0), I(0), R(0)) = (N - I(0), 500, 0)$, where $N = 10^6$ individuals. The black square data point represents the ideal seroprevalence data while the cyan circle represents the observed/measured seroprevalence.

#### S8 Varying Transmission Rate Only

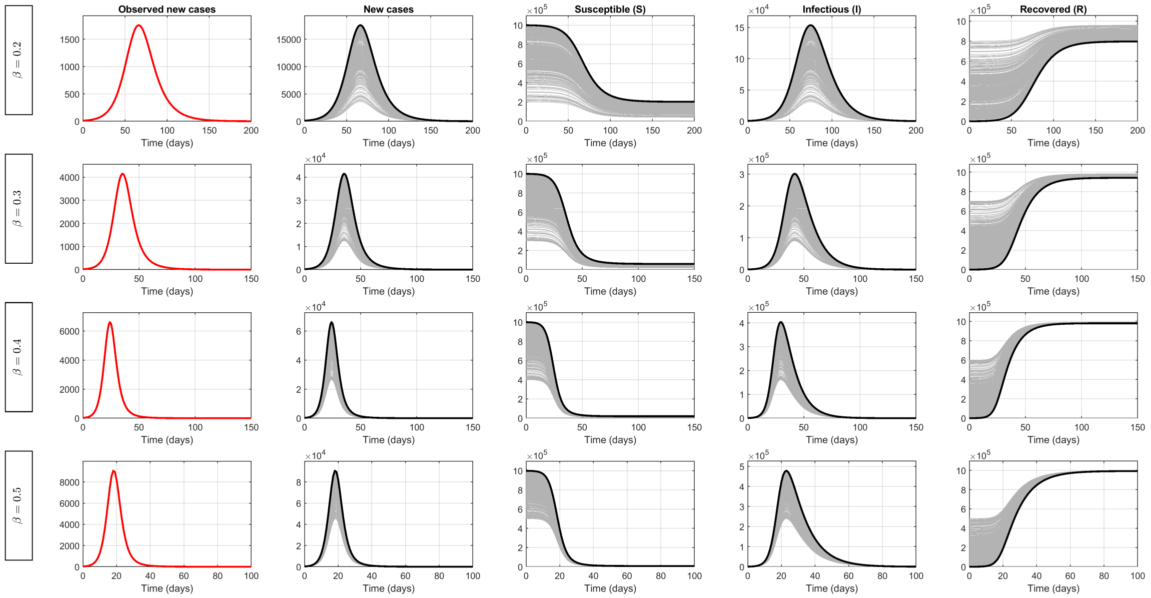

**Figure S9: The impact of varying transmission rate while fitting to detected new cases.** The first row is a replication of Figure 2B, while the remaining mimick Figure 2B while only varying transmission rate ($\beta$). Specifically, row one, two, three, and four correspond to $\beta$ values of 0.2, 0.3, 0.4, and 0.5 day$^{-1}$, respectively. The curves obtained through data fitting are depicted with a shaded gray color. The true dynamics, given with black curves, are generated with parameters $\gamma = 0.1$ day$^{-1}$, and initial conditions $(S(0), I(0), R(0)) = (N - I(0), 500, 0)$, where $N = 10^6$ individuals. Each row has a different number of parameter-fitted curves.

#### S9 Varying Case Ascertainment Ratio Only

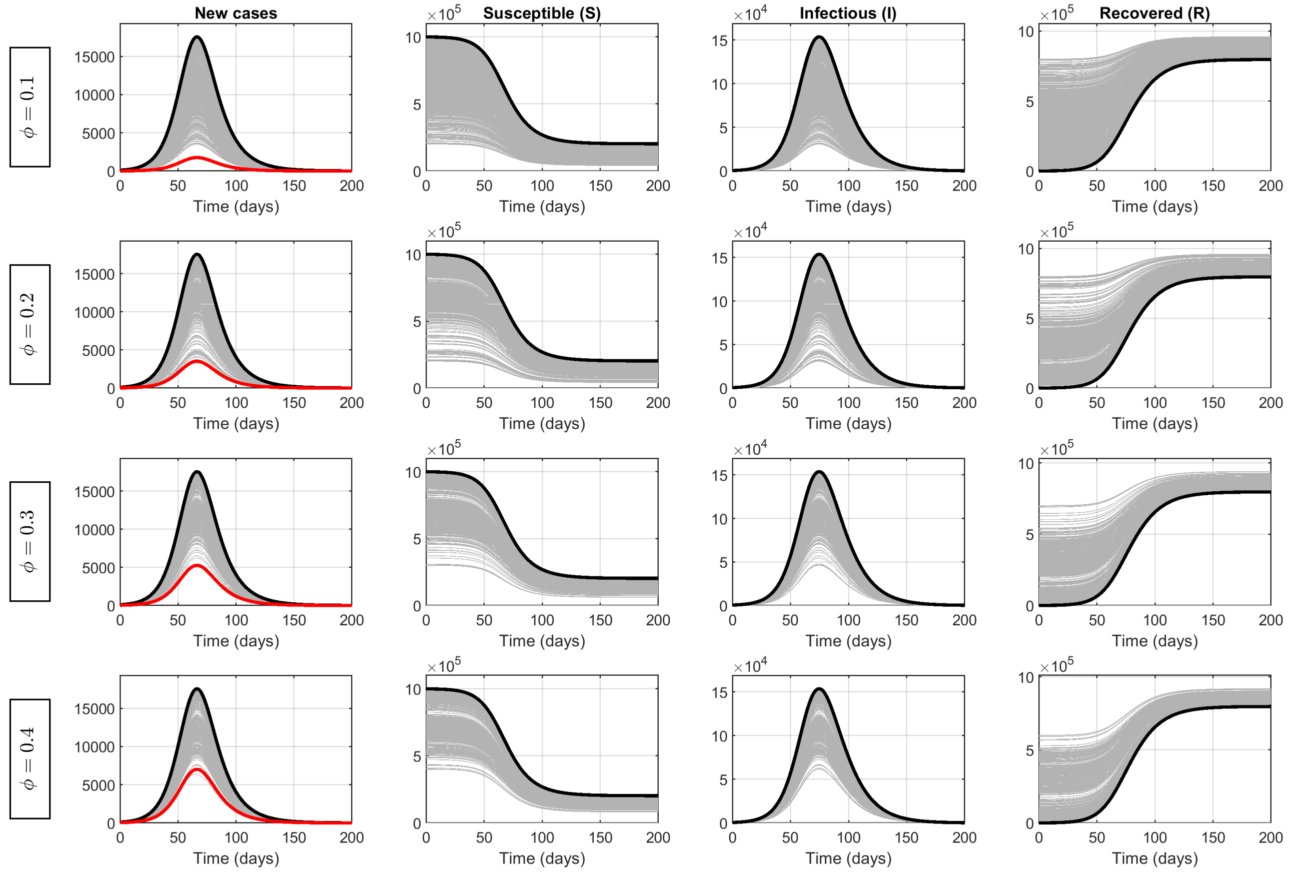

**Figure S10: Low case ascertainment ratio leads to larger uncertainties in QoIs.** The first row is a replication of Figure 2B, while the remaining mimick Figure 2B while only varying case ascertainment ratio ($\phi$). Row 1, 2, 3, and 4, correspond to $\phi$ value of $0.1, 0.2, 0.3$, and $0.4$. The curves obtained through data fitting are depicted with a shaded gray color. The true dynamics, given with black curves, are generated with parameters $\gamma = 0.1$ day$^{-1}$, and initial conditions $(S(0), I(0), R(0)) = (N - I(0), 500, 0)$, where $N = 10^6$ individuals.

#### S10 Varying Initial Number of Recovered Individuals

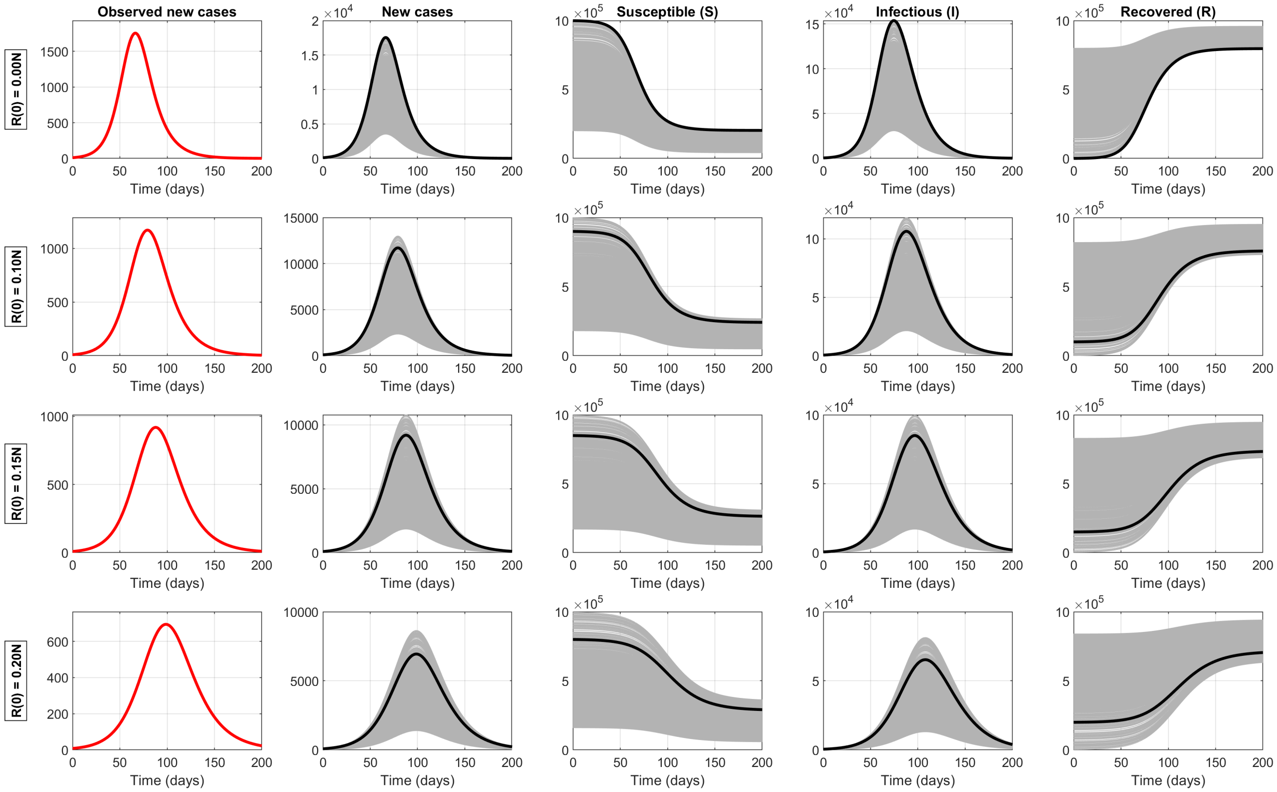

**Figure S11: Increasing initial number of immune/recovered individuals leads to trajectories on both sides of true new cases, S, I and R curves.** The first row is a replication of Figure 2B, while the remaining rows mimic Figure 2B while only varying the initial number of recovered individuals ($R(0)$). Rows 1, 2, 3, and 4 correspond to $R(0)$ values of 0, $0.1N$, $0.15N$, and $0.2N$, respectively. The curves obtained through data fitting are depicted in shaded gray, while the true dynamics are given by black curves. Unlike Figure 2, the fitted trajectories can be found on both sides of the true dynamics rather than being systematically biased. The true dynamics are generated with parameters $\beta = 0.2$ day$^{-1}$, $\gamma = 0.1$ day$^{-1}$, and initial conditions $(S(0), I(0), R(0)) = (N - I(0) - R(0), 500, R(0))$, where $N = 10^6$ individuals.

#### S11 Bayesian Inference on Noisy Data

In this section, we perform Bayesian inference of initial states $S(0)$, $I(0)$, $R(0)$, epidemic parameters $\beta$ and $\gamma$, and observation variance parameters $\sigma_I$ and $\sigma_S$ by fitting synthetic SIR model data with varying noise levels. The analysis is organized as follows. Section S11.1, S11.2, and S11.3 present inference results when fitting (i) only seroprevalence data, (ii) only newly detected cases, and (iii) both data streams simultaneously, respectively. All inferences use the No-U-Turn Sampler (NUTS) MCMC method.

The prior distributions for initial states $S(0)$, $I(0)$, $R(0)$, epidemic parameters $\beta$ and $\gamma$, and observation variance parameters $\sigma_I$ and $\sigma_S$ are shown in Figure S12. In all figures, the red cross indicates the true parameter value. These true values remain constant across all parameters, initial conditions, and observation variances for all simulations in this section—with the sole exception of $\sigma_I$, which varies according to the simulated noise level. Specifically, noise levels of approximately 10% and 40% correspond to $\sigma_I$ values of 0.1 and 0.4, respectively. The subsection on fitting to seroprevalence data only considered approximately 10% noise in seroprevalence data, denoted by $\sigma_S = 0.1$. Meanwhile, for the other two subsections, where we fit (i) only detected incidence data and (ii) both seroprevalence and detected incidence data, we consider 10% and 40% observation noise in detected incidence data (denoted by $\sigma_I$). For all these subsections, we produce two separate plots showing (a) posterior distributions and (b) trace plots.

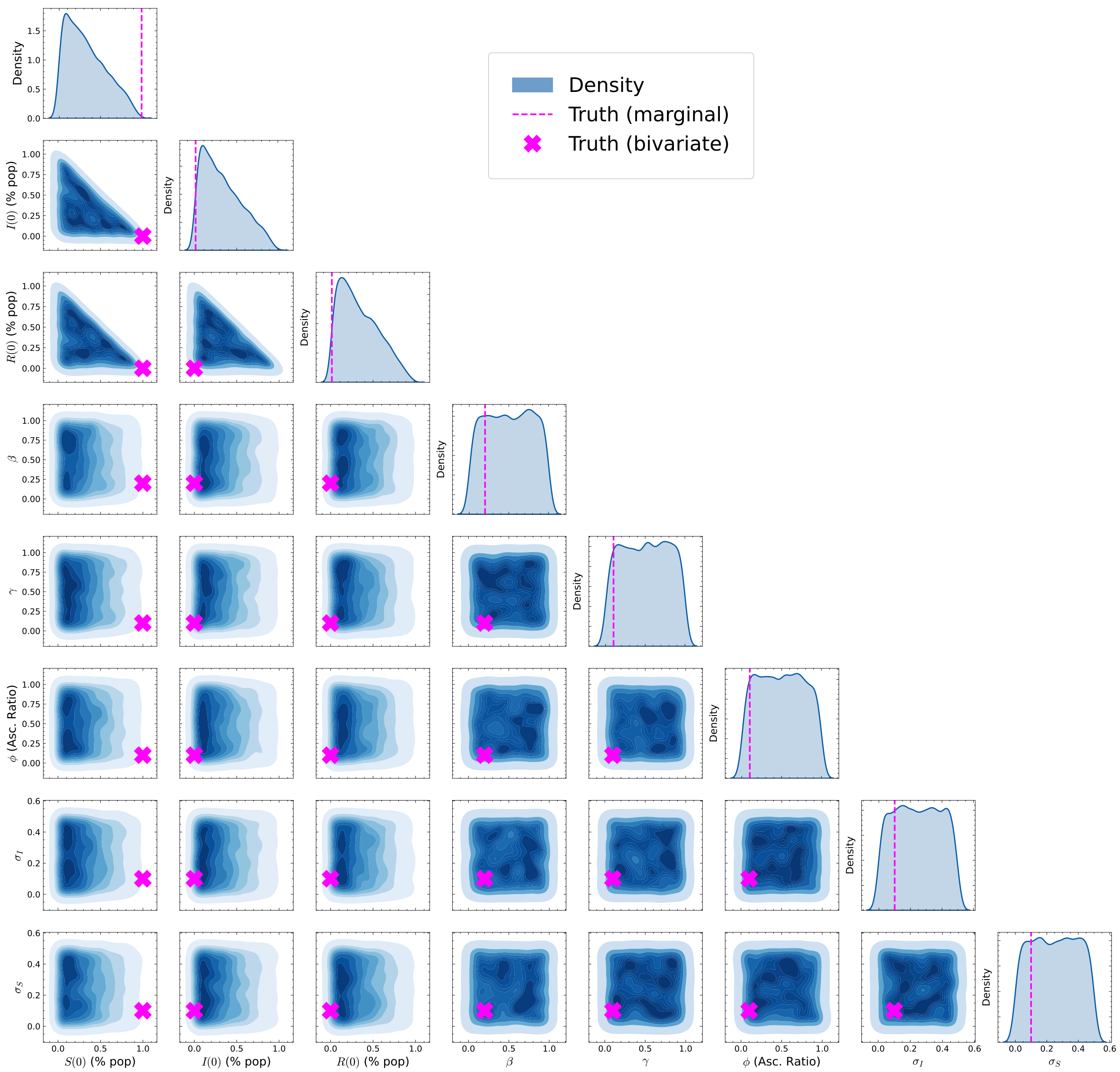

**Figure S12:** Prior distribution plots for an SIR epidemic model before MCMC fitting to synthetic data. The diagonal histograms show marginal probability densities for model parameters including initial states $S(0), I(0), R(0)$, epidemic parameters $\beta$ and $\gamma$, and observation variances $\sigma_I$ and $\sigma_S$. Off-diagonal entries show bivariate scatter plots showing joint prior distributions between pairs of parameters. The relatively wide distributions reflect initial uncertainty about parameter values before incorporating empirical evidence. The red cross indicates the true parameter value. Apart from the noise parameter $\sigma_I$, the true value for all parameters, initial condition, and noise parameter $\sigma_S$ remains the same for the purpose of simulations carried out in this section. Specifically, noise levels of approximately 10% and 40% correspond, respectively, to $\sigma_I$ values of 0.1 and 0.4.

#### S11.1 Fitting to noisy seroprevalence data only with $\sigma_S = 0.1$

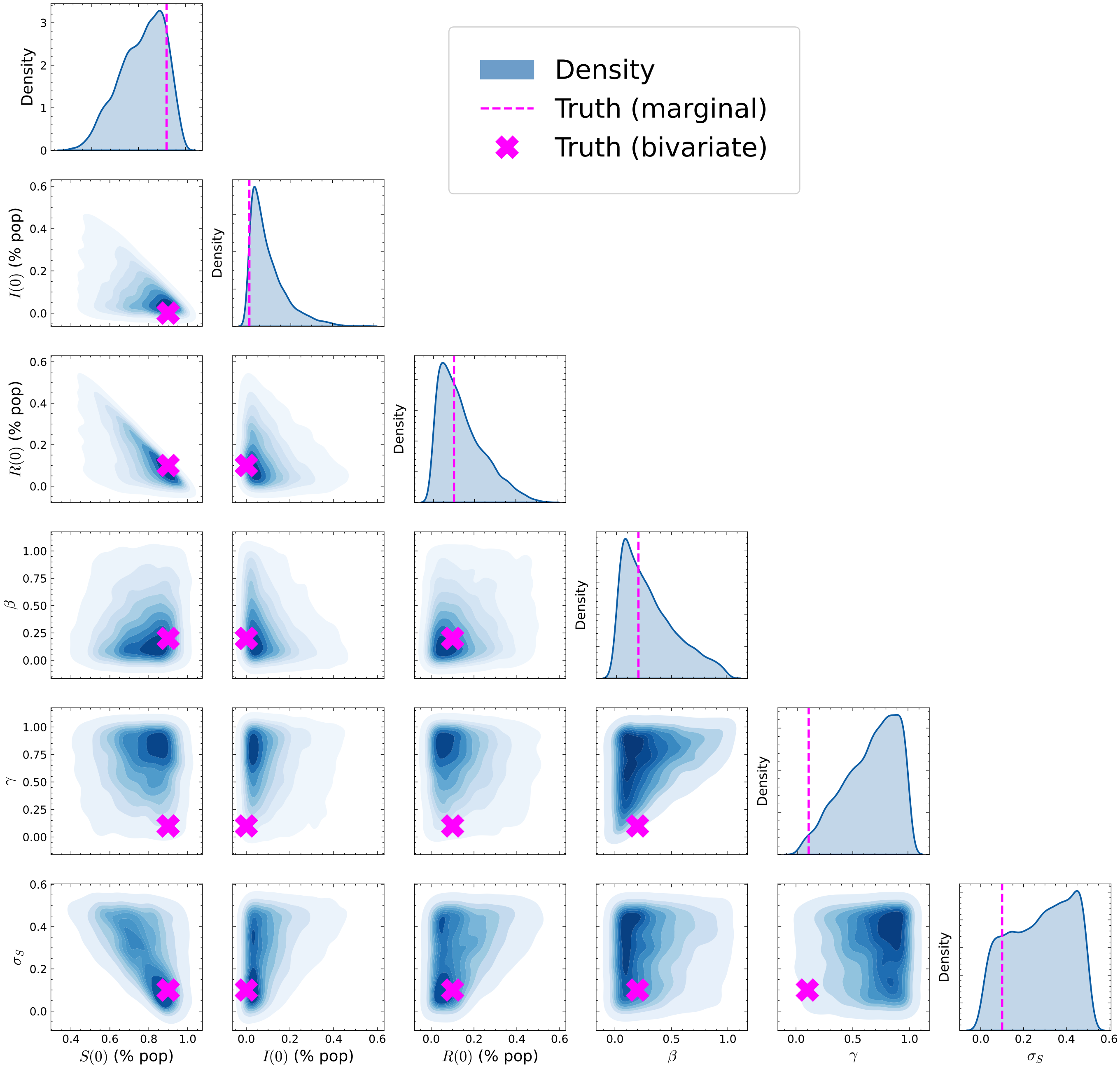

**Figure S13: Poor identifiability of epidemic parameters with wide exploration of parameter space, suggesting a single seroprevalence data provides insufficient information for precise parameter estimation.** Posterior distribution plots from an SIR epidemic model MCMC analysis. The diagonal shows marginal probability densities for each parameter, including initial population states $S(0), I(0), R(0)$, and model parameters $\beta$, $\gamma$, and observation variances $\sigma_I$ and $\sigma_S$. Off-diagonal elements display bivariate relationships between parameter pairs using kernel density estimation (KDE), revealing correlations in the posterior distribution.

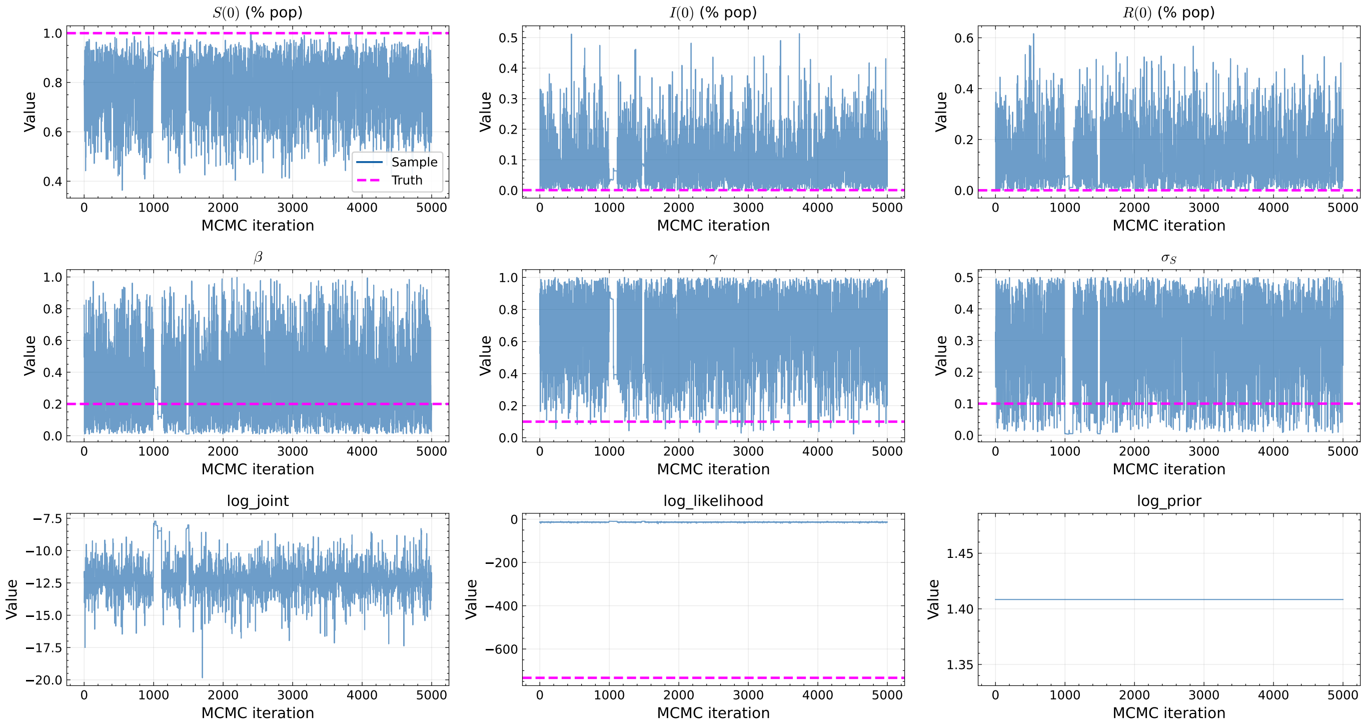

**Figure S14:** MCMC trace plots showing parameter evolution over 1,000 iterations when fitting the SIR model to seroprevalence data only. Parameter trajectories (blue lines) include initial population states ($S(0)$, $I(0)$, $R(0)$), epidemic parameters ($\beta$, $\gamma$, $\phi$), and observation variance parameters ($\sigma_I$, $\sigma_S$). True parameter values are indicated by magenta dashed lines. The bottom rows display log joint probability, log likelihood, and log prior probability traces.

#### S11.2 Fitting to noisy detected incidence data only

##### S11.2.1 $\sigma_I = 0.1$ and $\sigma_S = 0.1$

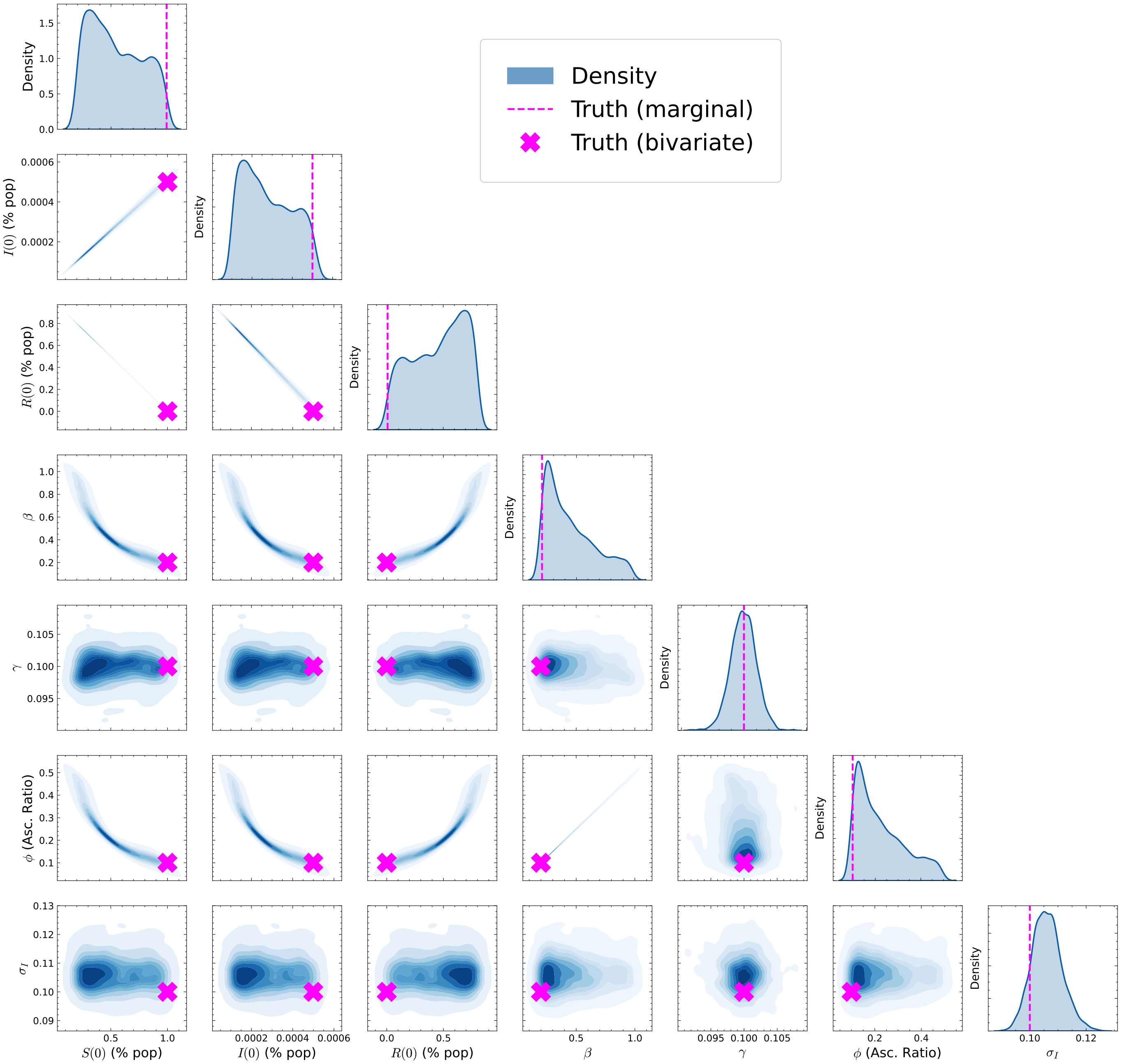

**Figure S15: Improved parameter identification compared to seroprevalence alone, though still showing substantial uncertainty in initial conditions and epidemic parameters.** Posterior distribution plots from an SIR epidemic model MCMC analysis. The diagonal shows marginal probability densities for each parameter, including initial population states $S(0), I(0), R(0)$, and model parameters $\beta$ (transmission rate), $\gamma$ (recovery rate), and observation variances $\sigma_I$ and $\sigma_S$. Off-diagonal elements display bivariate relationships between parameter pairs, revealing correlations in the posterior distribution. The concentrated distributions indicate reduced uncertainty after incorporating case data through Bayesian inference.

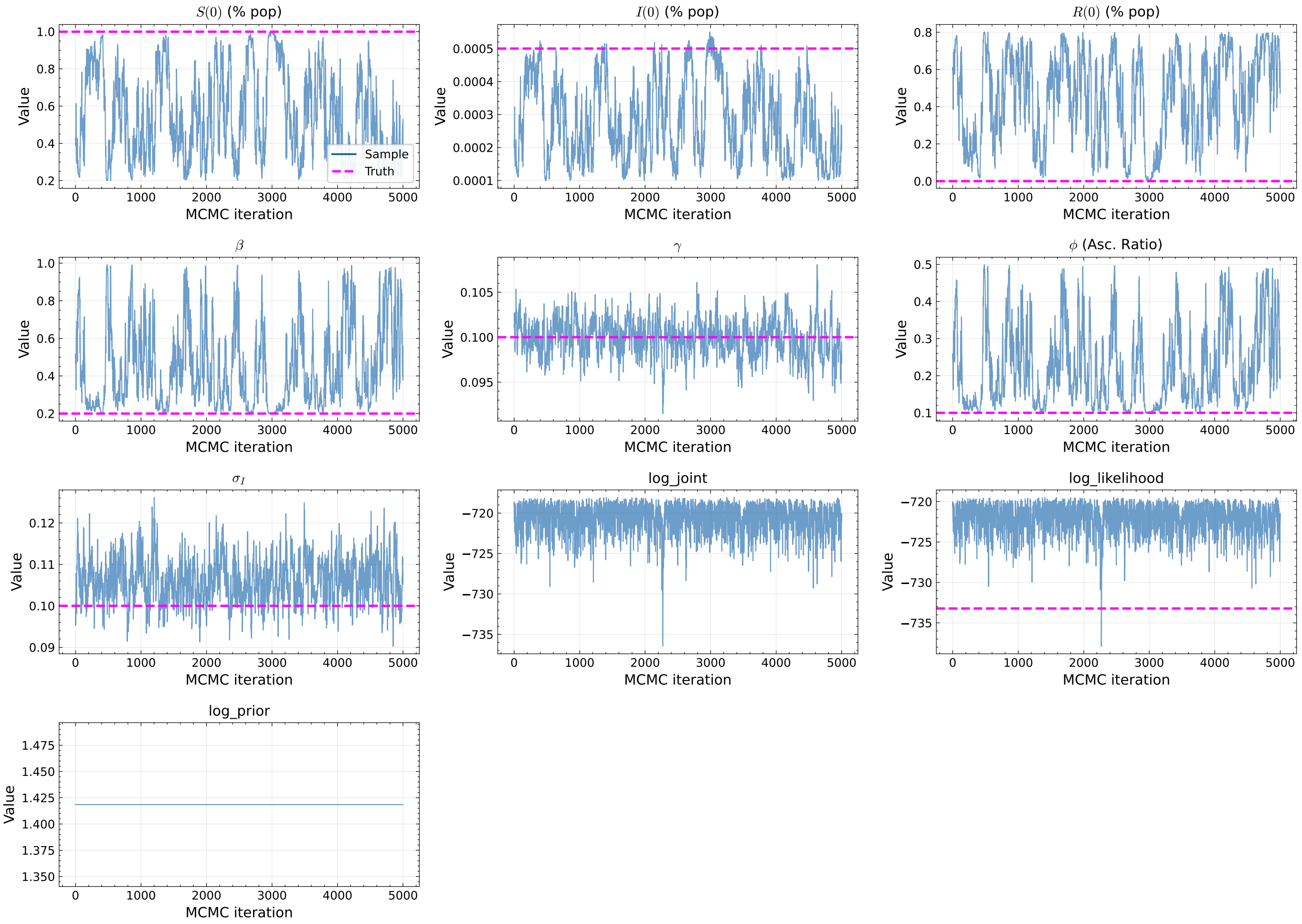

**Figure S16: MCMC trace plots showing parameter evolution over 1,000 iterations for an SIR epidemic model.** The top section displays time series for model parameters including initial population states ($S(0)$, $I(0)$, $R(0)$), epidemic dynamics parameters ($\beta$, $\gamma$, $\phi$), and observation model variances ($\sigma_I$, $\sigma_S$). Each parameter's sampling trajectory (blue line) is plotted against its true value (magenta dashed line). The bottom panels show the log joint probability, log likelihood, and log prior probability trajectories, indicating the model's fit quality throughout the MCMC sampling process. The significant fluctuations in parameter values across iterations demonstrate the algorithm's exploration of parameter space, while comparing sampled values with ground truth reveals the inference accuracy. These traces help diagnose MCMC convergence and mixing properties, showing whether the algorithm has adequately explored the posterior distribution of the epidemic model parameters.

##### S11.2.2 $\sigma_I = 0.4$ and $\sigma_S = 0.1$

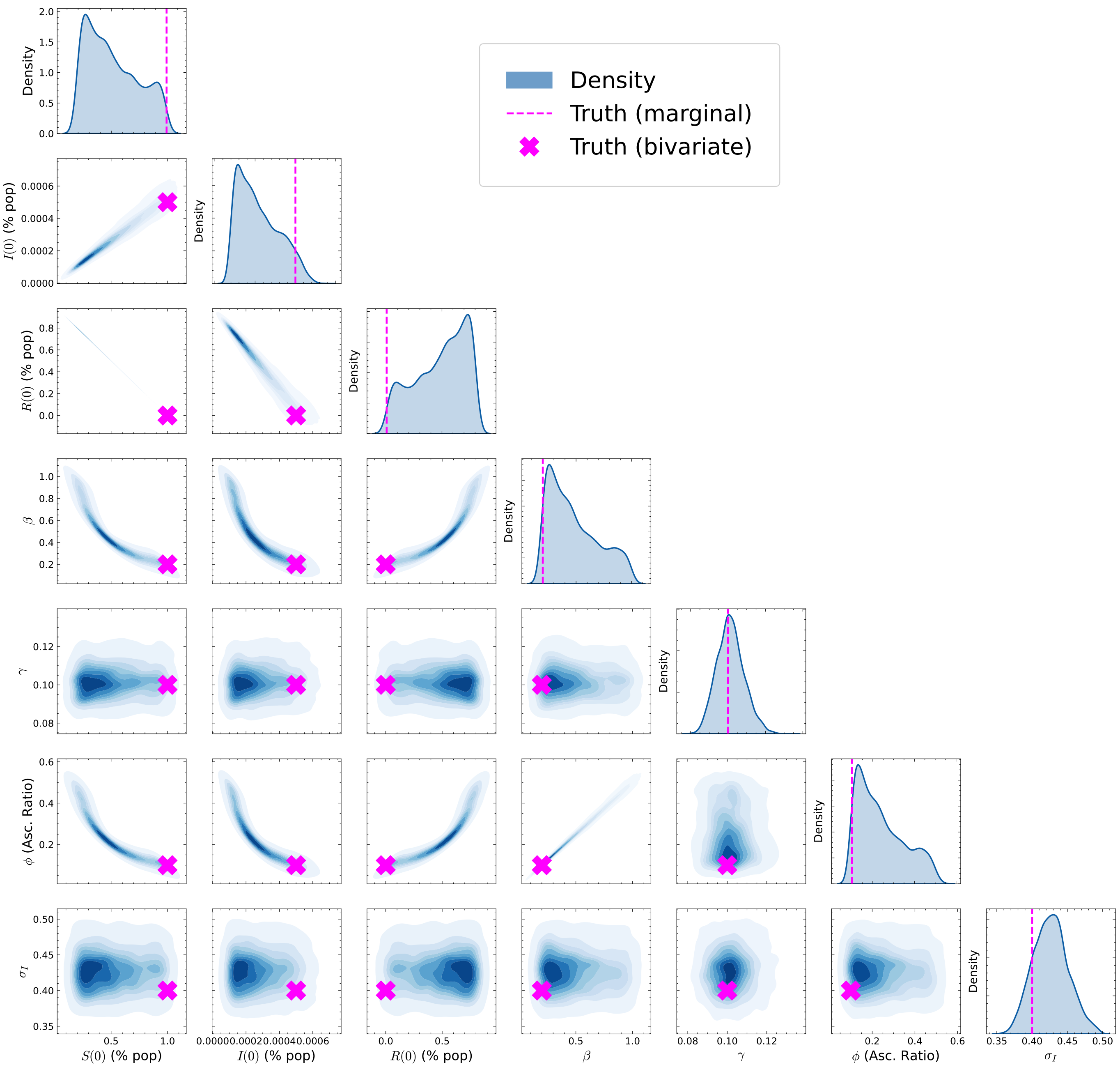

**Figure S17: Improved parameter identification compared to seroprevalence alone, though still showing substantial uncertainty in initial conditions and moderate uncertainty in epidemic parameters.** Posterior distribution plots from an SIR epidemic model MCMC analysis. The diagonal shows marginal probability densities for each parameter, including initial population states $S(0), I(0), R(0)$, and model parameters $\beta$, $\gamma$, and observation variances $\sigma_I$ and $\sigma_S$. Off-diagonal elements display bivariate relationships between parameter pairs using kernel density estimation (KDE), revealing correlations in the posterior distribution.

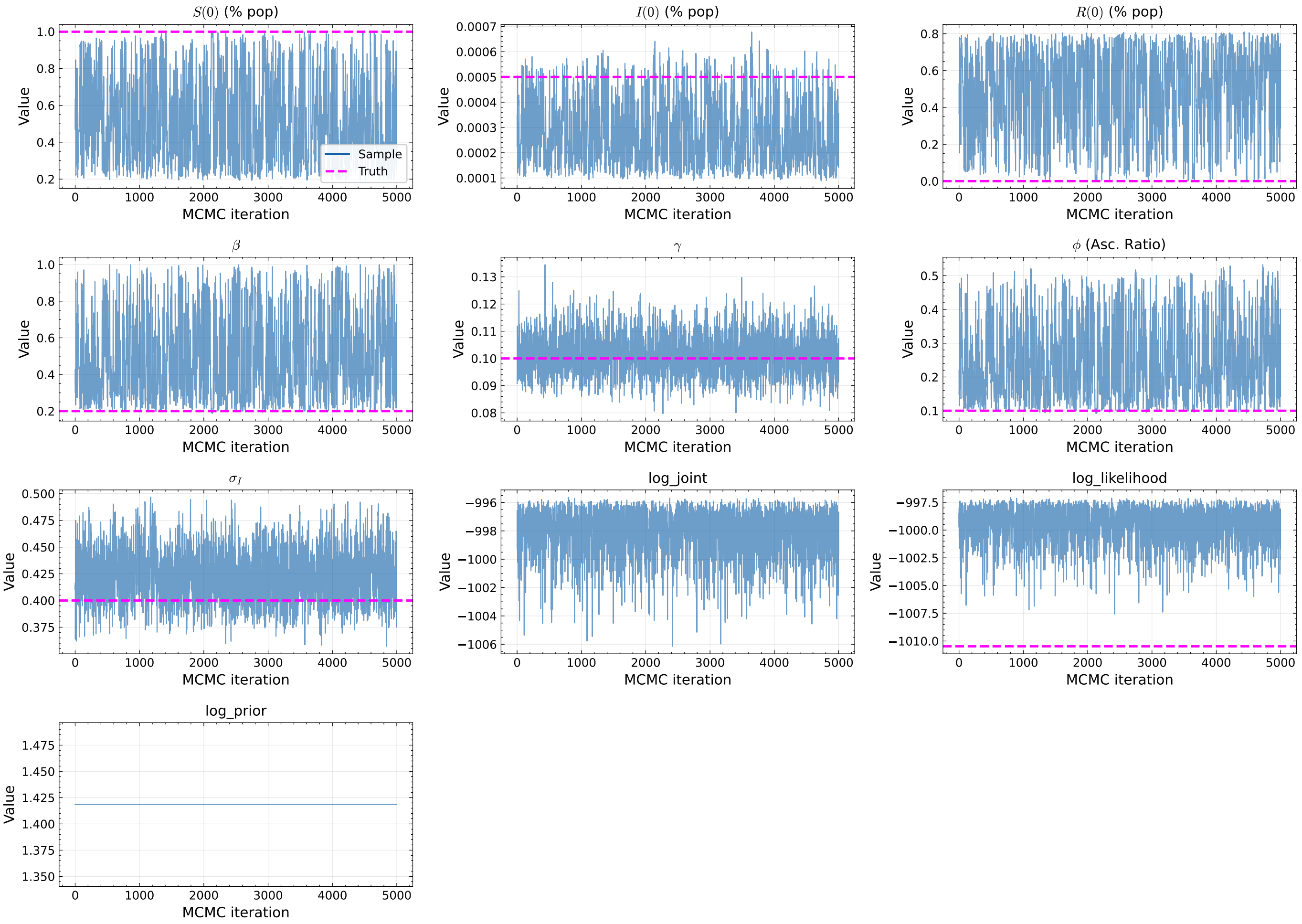

**Figure S18:** MCMC trace plots showing parameter evolution over 1,000 iterations when fitting the SIR model to seroprevalence data only. Parameter trajectories (blue lines) include initial population states ($S(0)$, $I(0)$, $R(0)$), epidemic parameters ($\beta$, $\gamma$, $\phi$), and observation variance parameters ($\sigma_I$, $\sigma_S$). True parameter values are indicated by magenta dashed lines. The last three subfigures display log joint probability, log likelihood, and log prior probability traces.

##### S11.3 Simultaneously fitting to noisy seroprevalence and detected incidence data

###### S11.3.1 $\sigma_I = 0.1$ and $\sigma_S = 0.1$

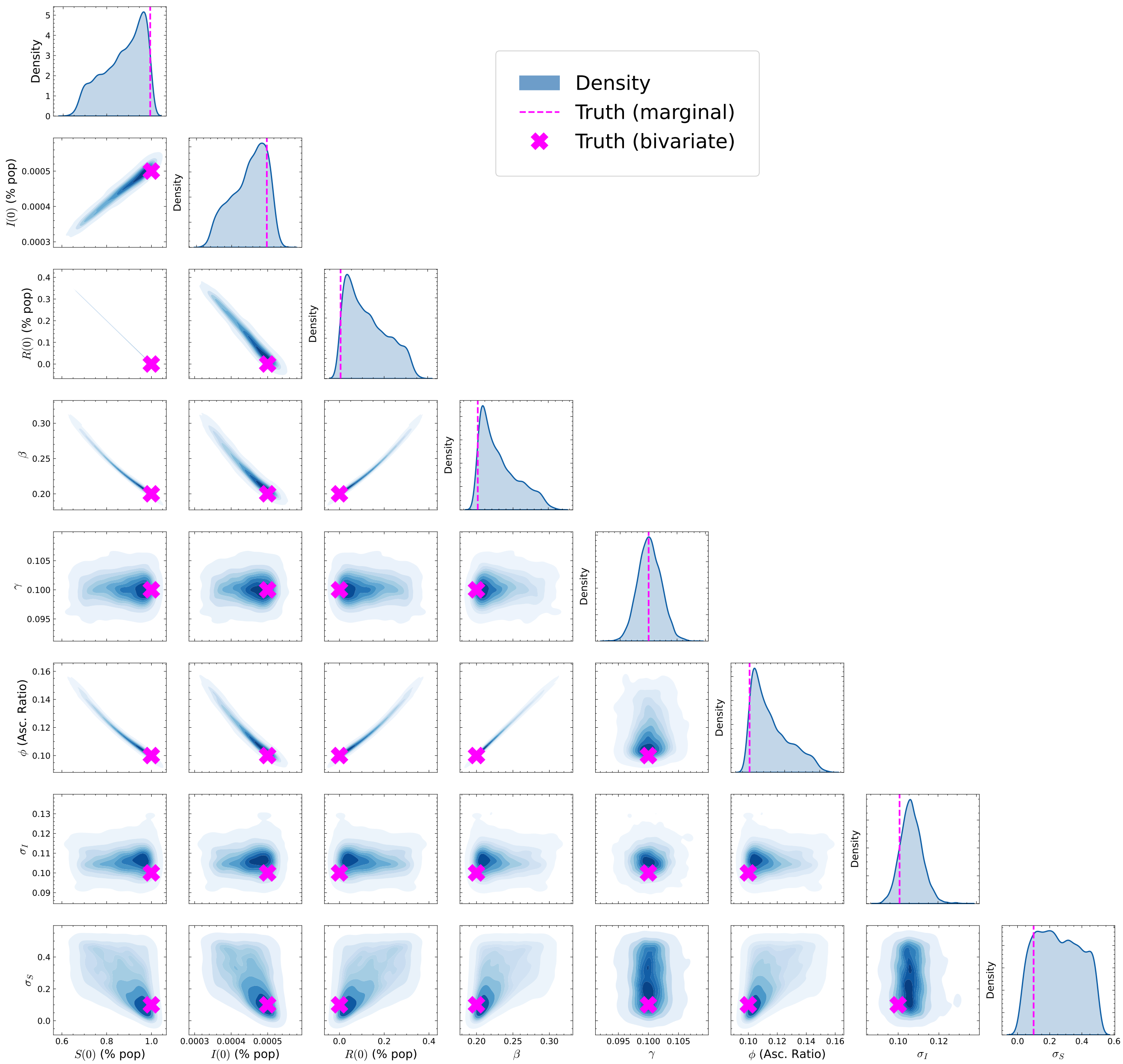

**Figure S19: Combining seroprevalence and case data substantially improves parameter identification, with narrower posterior distributions and more accurate estimates of true parameter values.** Posterior distribution plots from an SIR epidemic model MCMC analysis. The diagonal shows marginal probability densities for each parameter, including initial population states $S(0), I(0), R(0)$, and model parameters $\beta$, $\gamma$, and observation variances $\sigma_I$ and $\sigma_S$. Off-diagonal elements display bivariate relationships between parameter pairs using kernel density estimation (KDE), revealing correlations in the posterior distribution.

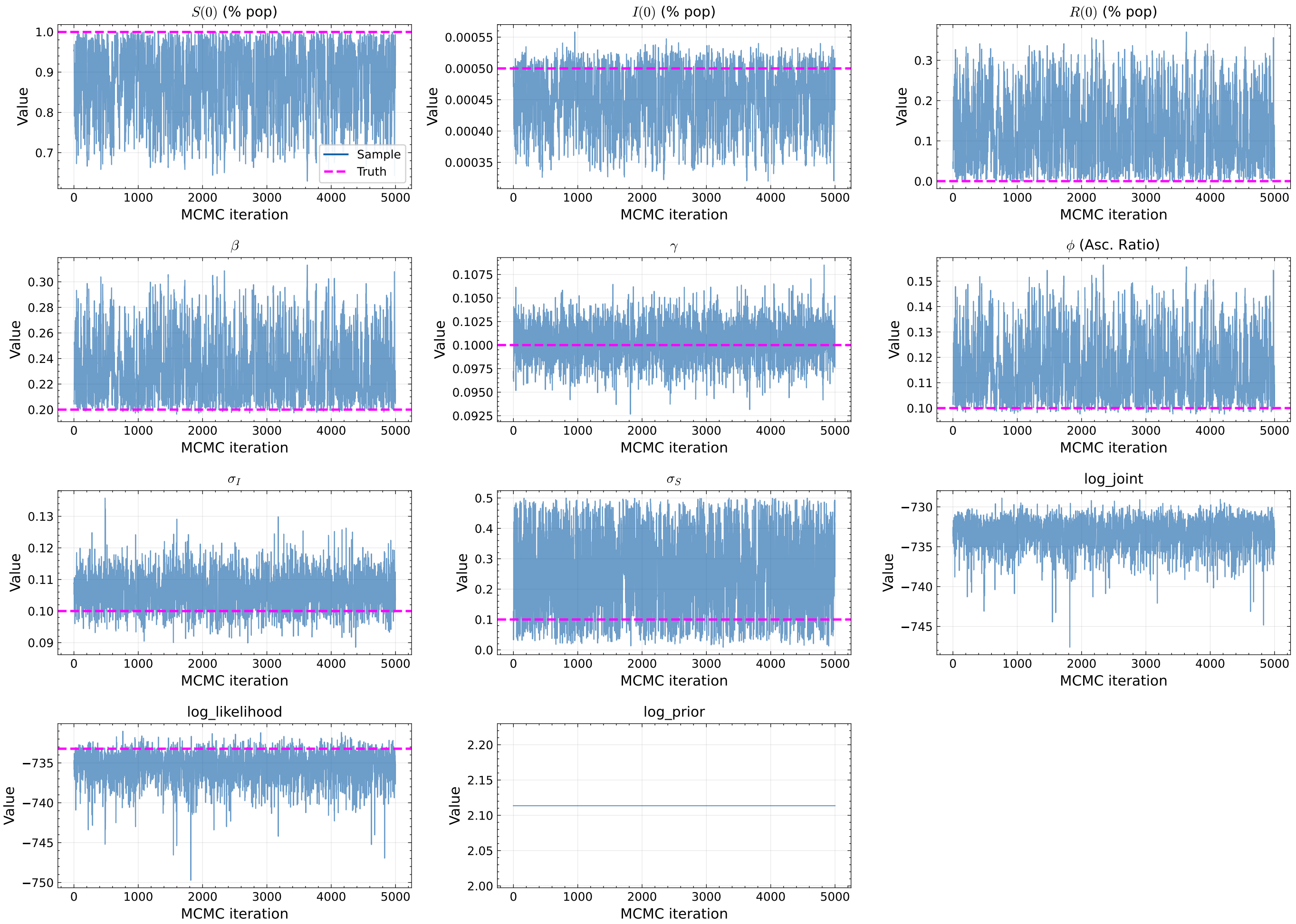

**Figure S20:** MCMC trace plots showing parameter evolution over 1,000 iterations for an SIR epidemic model. The top section displays time series for model parameters including initial population states ($S(0)$, $I(0)$, $R(0)$), epidemic dynamics parameters ($\beta$, $\gamma$, $\phi$), and observation model variances ($\sigma_I$, $\sigma_S$). Each parameter's sampling trajectory (blue line) is plotted against its true value (magenta dashed line). The bottom panels show the log joint probability, log likelihood, and log prior probability trajectories, indicating the model's fit quality throughout the MCMC sampling process. The significant fluctuations in parameter values across iterations demonstrate the algorithm's exploration of parameter space, while comparing sampled values with ground truth reveals the inference accuracy. These traces help diagnose MCMC convergence and mixing properties, showing whether the algorithm has adequately explored the posterior distribution of the epidemic model parameters.

##### S11.3.2 $\sigma_I = 0.4$ and $\sigma_S = 0.1$

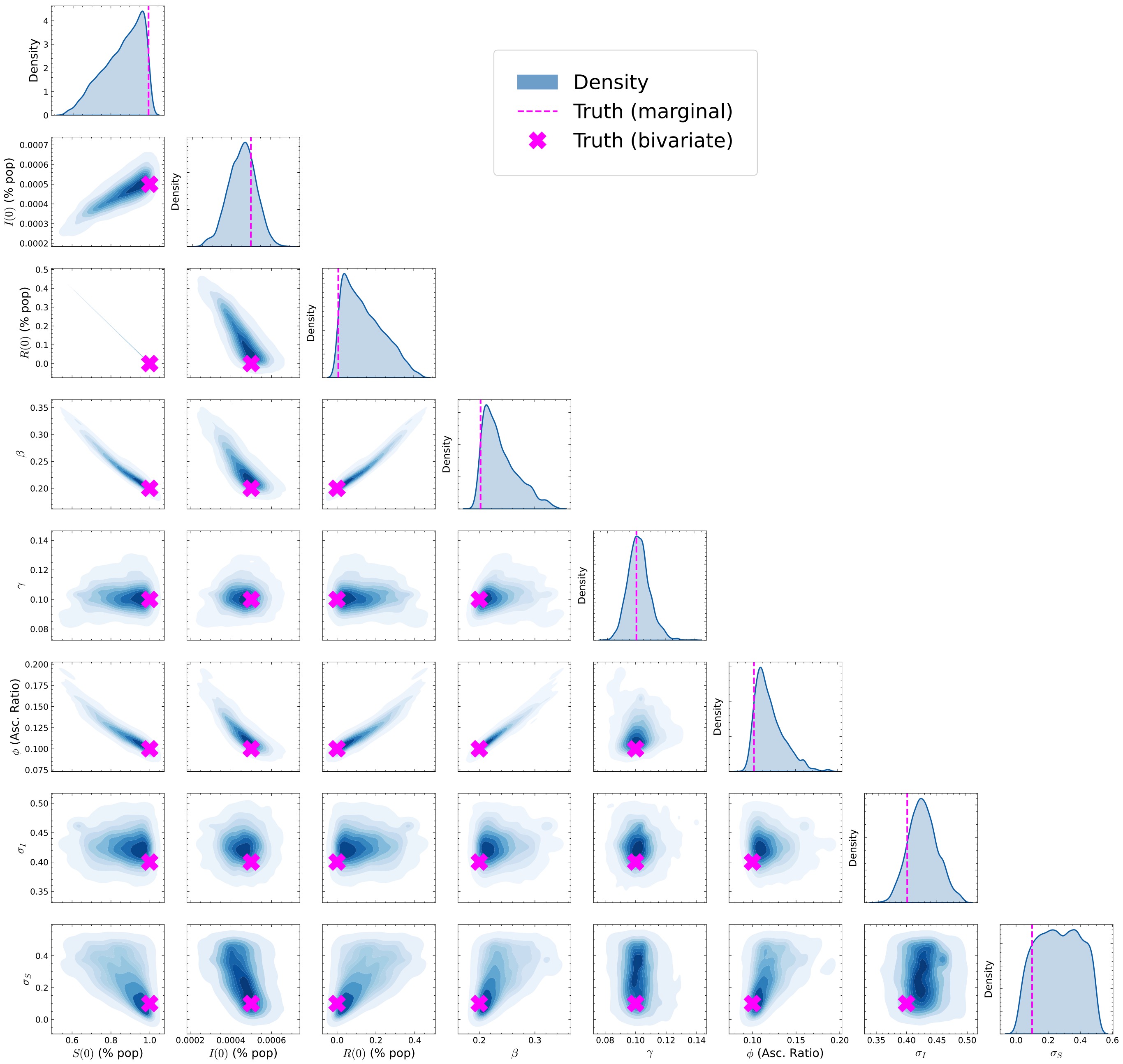

**Figure S21: Despite high observation noise, the model achieves good parameter identification by leveraging complementary information from both data streams, demonstrating the value of multiple data sources.** Posterior distribution plots from an SIR epidemic model MCMC analysis. The diagonal shows marginal probability densities for each parameter, including initial population states $S(0), I(0), R(0)$, and model parameters $\beta$, $\gamma$, and observation variances $\sigma_I$ and $\sigma_S$. Off-diagonal elements display bivariate relationships between parameter pairs using kernel density estimation (KDE), revealing correlations in the posterior distribution.

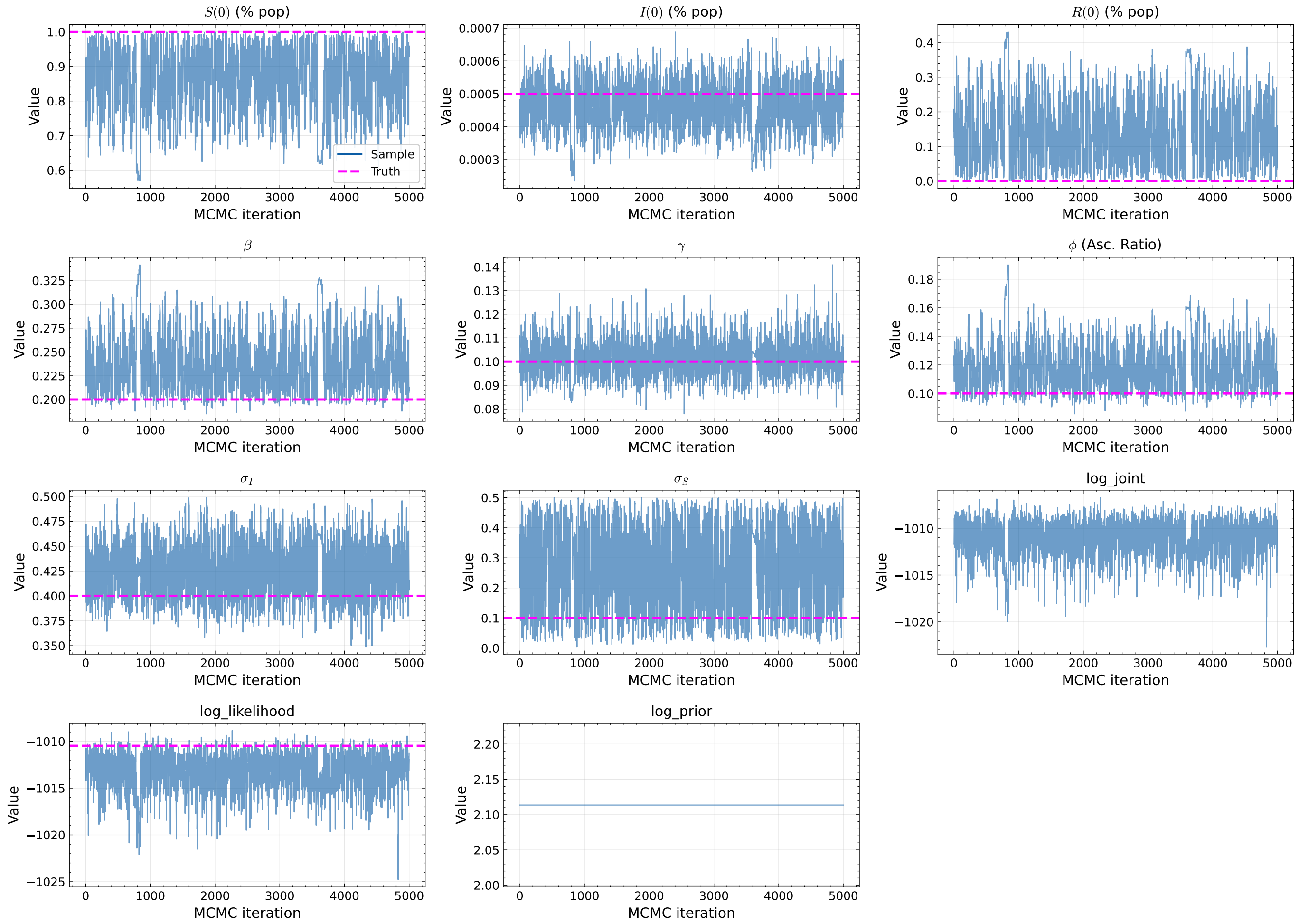

**Figure S22:** MCMC trace plots showing parameter evolution over 1,000 iterations for an SIR epidemic model. The top section displays time series for model parameters including initial population states ($S(0)$, $I(0)$, $R(0)$), epidemic dynamics parameters ($\beta$, $\gamma$, $\phi$), and observation model variances ($\sigma_I$, $\sigma_S$). Each parameter's sampling trajectory (blue line) is plotted against its true value (magenta dashed line). The bottom panels show the log joint probability, log likelihood, and log prior probability trajectories, indicating the model's fit quality throughout the MCMC sampling process. The significant fluctuations in parameter values across iterations demonstrate the algorithm's exploration of parameter space, while comparing sampled values with ground truth reveals the inference accuracy. These traces help diagnose MCMC convergence and mixing properties, showing whether the algorithm has adequately explored the posterior distribution of the epidemic model parameters.

#### S11.4 Uncertainty of effective reproduction number

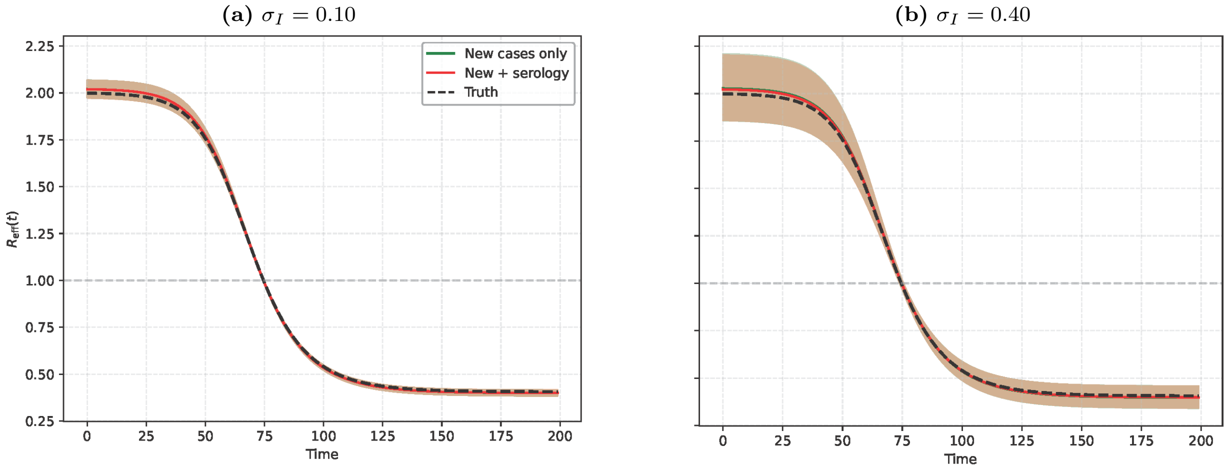

**Figure S23: The effective reproduction number maintains low uncertainty from noisy detected incidence alone.** Bayesian MCMC inference results showing the effective reproduction number with 95% credible intervals under different noise scenarios. **(a)** With 10% noise in detected incidence ($\sigma_I = 0.10$), fitting to new cases only (green line with narrow tan shading) produces tight credible intervals that accurately track true dynamics (black dashed line). Adding a single seroprevalence measurement (red line with shading) provides minimal improvement to the already low uncertainty from case data alone. **(b)** With 40% noise in detected incidence ($\sigma_I = 0.40$), fitting to new cases only (green line with wider tan shading) still maintains reasonable uncertainty bands that track true $\mathcal{R}_e(t)$, though credible intervals widen compared to the low-noise scenario. Combining with seroprevalence data (red line with narrower shading) modestly reduces uncertainty. The horizontal gray dashed line marks the critical epidemic threshold $\mathcal{R}_e(t) = 1$. These results demonstrate that $\mathcal{R}_e(t)$ exhibits relatively low uncertainty even from detected incidence data alone, consistent with its structural identifiability established in Supplementary Section S1.3, and that adding seroprevalence data does not dramatically reduce the already low uncertainty from case data alone.
